# Deep-Tissue Hemodynamic Sensing: Comparing Impedance and Photoplethysmography for Wearable Blood Pressure Estimation

**DOI:** 10.64898/2026.06.17.26355894

**Authors:** Seamus Thomson, Seobin Jung, Alexandros Pantelopoulos, Aniket Deshpande, Lawrence Cai, Emily Blanchard, Jackie Wasson, Debanjan Mukherjee, Lindsey Sunden, Sam Sheng, Shwetak Patel

**Author notes:** Co-First Authors (Seamus Thomson, Seobin Jung). Work done while at Google (Alexandros Pantelopoulos, Jackie Wasson).

## Abstract

The pursuit of continuous, cuffless blood pressure (BP) monitoring is constrained by the superficial sensing depth of photoplethysmography (PPG). Impedance plethysmography (IPG) offers deeper tissue penetration, but its comparative value over PPG remains unquantified at scale. In this comparative study of 261 participants (130 hypertensive, 131 non-hypertensive), we utilized a custom dual-modality wearable prototype to capture simultaneous IPG and PPG signals. Over 150,000 cardiac cycles were analyzed using an unsupervised archetype discovery pipeline to quantify beat-to-beat morphological heterogeneity. IPG resolved up to three distinct morphological modes per participant, whereas co-located PPG converged into highly conserved, uniform profiles. IPG captured specific signatures of pathological arterial remodeling and physiological habitus; ventral forearm IPG pulse amplitude exhibited a significant main effect for BP status (p = 0.024), a relationship absent in the co-located PPG signal. Furthermore, increasing body mass index (BMI) significantly attenuated the prevalence of steep-upstroke archetypes in IPG (p = 0.035), quantifying a likely damping effect of adipose tissue. Deep-tissue bioimpedance captures rich, heterogeneous hemodynamic signatures including arterial-dominant morphologies that are invisible to optical sensors. Transitioning from optical pulse wave analysis to bioimpedance-based models may offer a promising pathway for accurate wearable cardiovascular monitoring.

## 1 Introduction

The continuous, non-invasive monitoring of blood pressure (BP) remains a paramount unmet need in cardiovascular health management [1]. While intra-arterial catheterization (A-line monitoring) serves as the clinical gold standard for observing continuous pressure fluctuations, it also reveals that the hemodynamic waveform undergoes distinct transformations as it propagates across the arterial tree [18] (Fig. 1A). Pathological remodeling associated with hypertension is known to induce specific morphological shifts in these waveforms, effectively encoding vital biomarkers of cardiovascular risk [17] (Fig. 1B). However, the invasive nature of A-line confines its use to high-acuity settings, leaving a critical gap in our ability to track these hemodynamic biomarkers in daily life.

**Figure 1:**
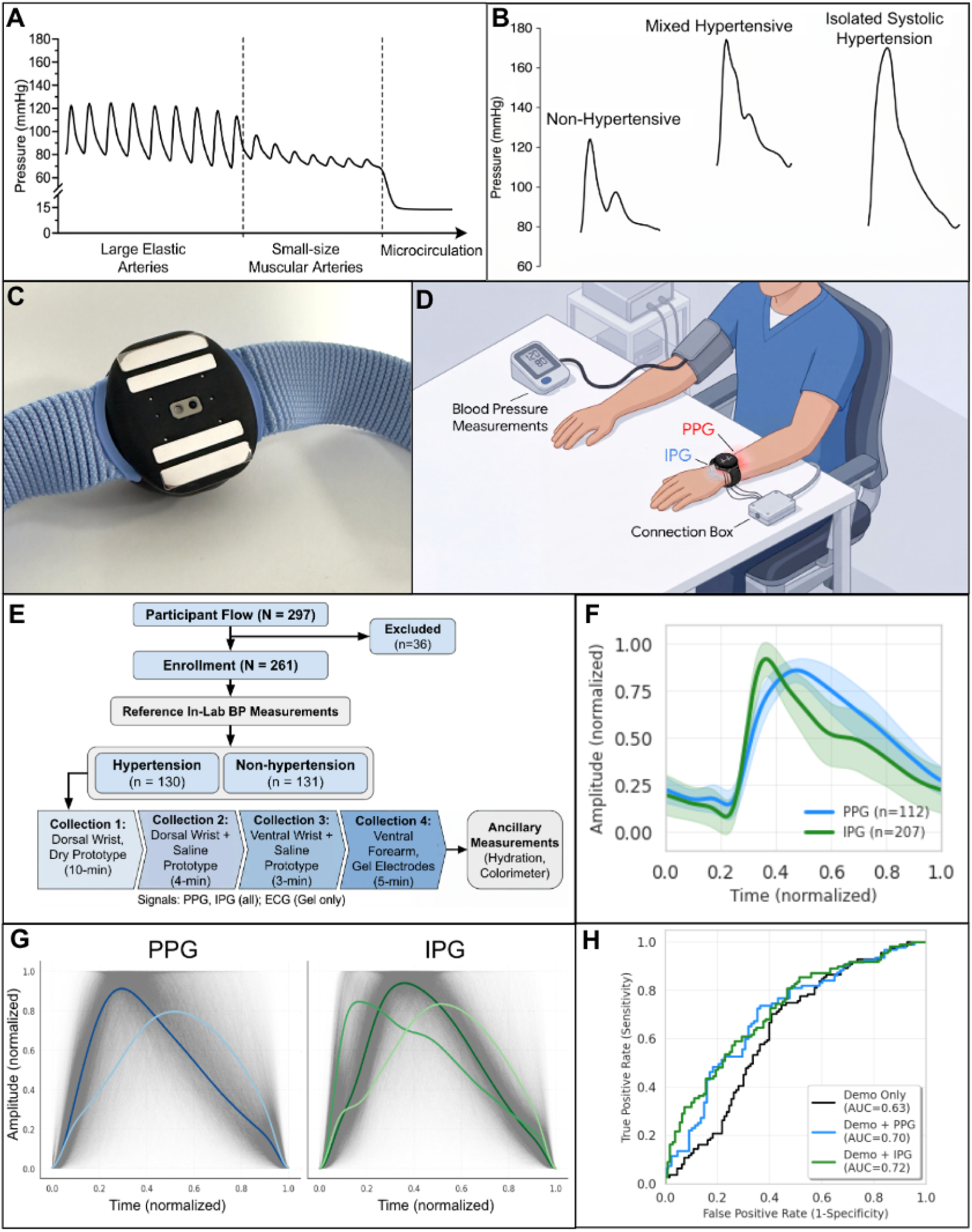
Deep-tissue hemodynamic sensing and evaluation framework. (A) Evolution of the arterial pressure waveform as it propagates from proximal elastic arteries to peripheral microvasculature. (B) Distinct morphological shifts in arterial line waveforms induced by hypertensive remodeling (adapted from [17]). (C) Photograph of the custom-engineered dual-modality wearable prototype featuring a four-electrode IPG array and a co-located infrared PPG module. (D) Experimental setup depicting in-lab reference blood pressure measurements and dual-modality waveform acquisition. (E) Study enrollment and data collection flowchart. (F) Comparison of aggregated, amplitude-normalized IPG and PPG pulsatile waveforms (mean ± 1 standard deviation) at the ventral forearm reference site. (G) Unsupervised K-means clustering of PCA-reduced beats at the dorsal wrist, highlighting the morphological heterogeneity captured by IPG (three distinct modes) compared to the unimodal convergence of PPG (two nearly identical modes). (H) Cross-validated Receiver Operating Characteristic (ROC) curves demonstrating the incremental predictive value of IPG-derived individual archetypes and PPG-derived waveform templates for hypertension classification over demographic baselines.

To circumvent these limitations, photoplethysmography (PPG) has been widely explored in consumer wearables for cuffless BP estimation [10, 30]. However, the robust translation of PPG into reliable hemodynamic biomarkers is fundamentally constrained by its shallow optical sensing depth [2]. Because PPG relies on light absorption within the superficial dermal microvasculature, the resulting waveforms capture downstream consequence rather than the primary arterial pressure wave itself. Consequently, PPG signals are heavily confounded by localized skin-level factors, including ambient temperature, sensor contact pressure, skin tone, and peripheral vasoconstriction [2, 3] and are frequently decoupling it from central hemodynamic changes [9].

Impedance plethysmography (IPG) offers a promising complementary or alternative approach to bypass these superficial limitations of optical sensing [4, 5, 6]. By injecting an imperceptible, high-frequency alternating current through tissue, IPG penetrates beyond the hypodermis to interrogate the deeper conductive volume of the limb. This superior sensing depth could be an essential differentiator for wearable blood pressure estimation. While chronic hypertension induces structural vascular remodeling - such as increased arterial stiffness and medial hypertrophy [8] - these pathophysiological changes are disproportionately pronounced in small muscular arteries compared to large elastic arteries. Comparative clinical studies reveal a striking disparity: while large artery elasticity may decrease by approximately 19% in hypertensive individuals compared to non-hypertensive controls, the radial and ulnar arteries - the primary vessels interrogated by peripheral sensors - are classified as small arteries which exhibit an average 72% reduction in compliance during hypertension [11]. We hypothesize that by reaching these deeper vascular structures, IPG captures rich, beat-to-beat hemodynamic biomarkers and morphological heterogeneities that remain invisible to superficial optical sensors.

Despite these theoretical biophysical advantages, the incremental value of IPG over PPG has not been quantified at scale. This study addresses this gap by presenting a large-scale physiological discovery pipeline. Leveraging a custom dual-modality wearable prototype across a stratified cohort of 261 hypertensive and non-hypertensive individuals, we characterize the deep-tissue morphological heterogeneity unique to bioimpedance. By isolating these novel, IPG-specific hemodynamic biomarkers, we aim to demonstrate that moving beyond superficial optical sensing could be fruitful for the future of wearable cardiovascular monitoring.

## 2 Methodology

### 2.1 Study Design and Ethical Approval

This prospective, observational feasibility study was designed to quantify the comparative performance of IPG versus PPG for non-invasive BP estimation. The protocol was approved by the WCG Institutional Review Board (IRB, 20251519), conducted across sites in San Francisco and Mountain View, CA, and in accordance with the Declaration of Helsinki. All participants provided written informed consent prior to enrollment. To protect participant privacy, all study data were de-identified in accordance with IRB standards.

A cross-sectional observational approach was adopted to compare signal features between hypertensive and non-hypertensive cohorts, thereby avoiding the confounding heart rate (HR) shifts typical of interventional trials [15]. A statistical power analysis (G*Power; [13]) indicated that a sample size of 91 participants per group (182 total) was required to detect differences in four features using a two-tailed Welch’s t-test with a medium effect size (Cohen’s d = 0.5, *α*_corrected_ = 0.0125, power = 0.8). Our final analyzed cohort (N = 261) exceeded this minimum requirement.

### 2.2 Participant Cohort

Participants were recruited from internal and external sources (N = 261 enrolled from an eligible pool of N = 297). Inclusion criteria required participants to be at least 18 years of age and capable of providing informed consent. Exclusion criteria were strictly enforced to ensure both participant safety and signal integrity. Individuals were ineligible if they: 1) possessed internal electronic or metallic implants (e.g., pacemakers or defibrillators), as IPG introduces an active alternating current; 2) were currently taking diuretic or steroid medications, which alter fluid volume and vascular tone; or 3) were pregnant or suspected pregnancy, or 4) were unwilling or unable to provide informed consent.

Stratification into hypertensive (HTN_True, n = 130) and non-hypertensive (HTN_False, n = 131) cohorts was based on in-lab BP measurements obtained via automated arm cuffs (Welch Allyn / Omron). Following American Heart Association (AHA) guidelines, participants were assigned to the HTN_True group if their average Systolic Blood Pressure (SBP) was *≥* 130 mmHg or Diastolic Blood Pressure (DBP) was *≥* 80 mmHg. Participants falling below 130/80 mmHg were assigned to the HTN_False group. This grouping methodology mirrors the protocol established in the Apple Hypertension study [12]. Finally, the cohort was partitioned into a Training Set (n = 219) for feature discovery and an independent Hold-out Test Set (n = 42) for performance validation.

To account for physiological habitus, a known confounder for blood pressure and could potentially influence the bioimpedance conductive pathway, participant body mass index (BMI) was documented. Because adipose tissue possesses a lower electrical conductivity than lean muscle, variation in localized fat distribution introduces a potential mechanism for modulating the depth and volume of the current paths interrogated by IPG. Following the Centers for Disease Control and Prevention (CDC) guidelines, participants were categorized into four groups: Underweight (BMI *<* 18.5 kg/m^2^), Healthy Weight (18.5 - 24.9 kg/m^2^), Overweight (25.0 - 29.9 kg/m^2^), and Obese (BMI *≥* 30.0 kg/m^2^).

To prevent demographic or physiological confounding across our machine learning pipelines, data partitioning was structurally balanced using a multi-key stratification strategy based on BP groups, age bucket (18-39, 40-59, and 60+ years), and obesity status similar to the Apple Hypertension study. This cross-stratification ensured that the Training Set and the independent Hold-out Test Set maintained highly representative joint distributions of age, body habitus, and clinical diagnostic labels.

### 2.3 Dual-Modality Wearable Prototype and Sensor Data Acquisition

The study utilized a custom-engineered, wrist-worn prototype based on a modified commercial smartwatch form factor (Google Pixel Watch 2). The device integrated simultaneous IPG and PPG sensing configurations on its wrist-facing underside (Fig. 1C). Key design features, detailed previously by [7], include a 316L stainless steel four-electrode array for IPG with an 18 mm center-to-center sensing gap and a 0.25 mm protrusion to ensure consistent skin contact. An infrared reflective PPG module (OPB733TR) was co-located at the center of the IPG array. Analog signals were captured at 2 kHz using a Biopac MP36R system. All data underwent daily quality assurance by trained personnel to ensure signal integrity across the recruitment period.

### 2.4 Experimental Protocol

Participants were seated at a table with arms at heart level (Fig. 1D). Following an initial 5-minute rest period, reference BP was measured using an automated cuff (Welch Allyn / Omron); the average of two subsequent measurements obtained from the arm with the higher pressure served as the ground truth. Data acquisition followed four progressive phases (Fig. 1E):

- Collection 1: Dorsal Wrist (Dry Interface). The wearable prototype was secured to the dorsal wrist using a standard Pixel Watch Gradient Stretch band. A 10-minute recording was captured to evaluate the performance of the dry stainless-steel electrodes under typical consumer-wear conditions.
- Collection 2: Dorsal Wrist (Saline Interface). To isolate the influence of skin-electrode contact impedance, the dorsal wrist skin was prepared with two wipes of a sterile saline (0.9% NaCl) pad. A 4-minute recording was acquired for comparison against the dry-electrode measurements.
- Collection 3: Ventral Wrist (Saline Interface). The prototype was rotated to the ventral aspect of the wrist following identical saline skin preparation. This 3-minute collection aimed to assess sensor sensitivity over the radial and ulnar arteries in an anatomical region with reduced bone and connective tissue interference.
- Collection 4: Ventral Forearm (Reference Standard). A final 5-minute multimodal reference measurement was obtained from the ventral forearm using Biopac modules instead of the prototype. A concurrent Lead I ECG was recorded to provide a definitive temporal reference for the cardiac cycle. The localization methodology, and sensor placement and spacing were kept identical to a previously validated configuration [5] to ensure cross-study consistency.

Ancillary characteristics at three sensing sites (left index finger, left ventral forearm, and left dorsal forearm) were documented. Skin hydration was quantified using a Delfin Moisture MeterSC, with three measurements recorded and averaged for each location. Skin color measurements (individual typology angle, ITA) were performed using a Colorimeter CL 400 (Courage + Khazaka electronic GmbH, Germany).

To mitigate motion-induced artifacts and environmental noise, participants were mandated to maintain absolute stillness and silence during data acquisition. Sensing fidelity was monitored in real time by a trained proctor using the digital acquisition interface. If significant signal contamination, atypical kinetic interference, or an absence of identifiable pulsatile morphology was observed, the session was temporarily suspended to troubleshoot connection integrity or sensor misplacement. All acquired time-series data from the BIOPAC system were exported as CSV files for subsequent offline processing and analysis.

### 2.5 Preprocessing and Feature Extraction

The IPG and PPG time-series were first subjected to a zero-phase 8th-order Butterworth bandpass filter (0.5 - 10.0 Hz) to remove baseline wander and high-frequency instrumental noise. To suppress high-frequency noise amplification, signal derivatives were computed using Savitzky-Golay filters. Morphological fiducials - specifically the pulse onset, systolic peak, max slope (*u*-point), max 2nd time derivative (*a*-point), and min 2nd time derivative (*b*-point) - were extracted using the pyppg software library [14]. These fiducials were utilized for all subsequent analysis. Finally, to facilitate cross-modality waveform comparison, individual cardiac cycles were segmented using onsets and *a*-points, linearly resampled, and amplitude-normalized to the range [0, 1].

### 2.6 Beat Aggregation using Cross-Correlation-based Signal Quality Check

To facilitate group-wise comparisons between the hypertensive and non-hypertensive cohorts, a high-fidelity representative heartbeat template was generated for each participant per measurement site. Aggregating individual cycles into a singular template minimized stochastic beat-to-beat noise and ensured that downstream morphological analyses were executed on stable, reproducible physiological representations.

High-quality beats were identified using a dual-stage Signal Quality Index (SQI) filtering pipeline adapted from established PPG telemetry frameworks [16]:

- Rule-based Feasibility Gate: Segments were first passed through a physiological filter to exclude beats with implausible heart rates (below 40 or above 180 bpm) or extreme inter-beat interval (IBI) variability (defined as max peak-to-peak interval*/*min peak-to-peak interval *≥* 2.2).
- Template Matching Algorithm: The remaining viable segments were evaluated using a sliding-window correlation approach. A local template was generated by averaging beats within a 10-second window. Individual cycles were retained for final aggregation only if they exhibited a mean Pearson correlation coefficient (*r*) *≥* 0.80 with this local template.

This SQI pipeline was applied across a uniform temporal window at the end of each recording block to capture stable, resting-state hemodynamics. Specifically, the final four minutes of data were analyzed for Collections 1, 2, and 4, while the final three minutes were utilized for Collection 3 due to its shorter stabilization period. Analysis windows were 10 seconds in duration with a 50% temporal overlap. All high-quality beats surviving both gates underwent linear tilt correction to eliminate baseline wander before being ensemble-averaged into a singular, high-fidelity representative template for each participant, sensor modality, and anatomical site. Both unnormalized templates and templates normalized in both time and amplitude are used throughout the visualization and data analysis.

### 2.7 Unsupervised Archetype Discovery using Clustering

During the evaluation of beat-aggregated waveforms, we observed pronounced morphological heterogeneity - the presence of multiple, distinct waveform modes within a single recording that a single ensemble average would inadvertently obscure. To isolate and resolve these discrete physiological variations without introducing prior bias, we developed an unsupervised archetype discovery pipeline executed via Principal Component Analysis (PCA) and *K*-means clustering.

Prior to dimensionality reduction using PCA, individual resampled cardiac cycles underwent an artifact rejection process. Cycles with durations falling outside 50% - 150% of the participant’s median IBI were discarded, and a per-beat linear tilt correction was applied to eliminate residual baseline drift. To extract the most salient morphological features from the remaining 256-dimensional resampled temporal beats while minimizing computational noise, the data was standardized (min-max scaling) and transformed via PCA. We retained the number of principal components required to explain *≥* 95% of the total variance, projecting each beat into a low-dimensional manifold capturing essential pulsatile characteristics (e.g., systolic upstroke velocity and diastolic decay constants).

Unsupervised *K*-means clustering was subsequently performed on the PCA-reduced feature space. The optimal number of clusters (*k*) was determined dynamically by evaluating *k* in the range [2, 6] and selecting the configuration that maximized the Mean Silhouette Coefficient - a measure of cluster cohesion and separation. If the maximum Silhouette score failed to exceed a heuristic threshold (e.g., 0.25), the signal was classified as having a single dominant morphology (*k* = 1). Once cluster boundaries were established, global centroids (archetypes) were generated by averaging the temporal vectors within each cluster to visualize the representative pulse waves.

This pipeline was deployed in two distinct analytical approaches:

- Global Analysis: PCA and *K*-means clustering were applied to the entire pooled dataset to identify universal morphological archetypes across the stratified population. This allowed for the direct visualization of representative pulse waves and a comparison of their prevalence between the hypertensive and non-hypertensive groups.
- Per-Participant Analysis: PCA and *K*-means clustering were performed for each participant individually to evaluate intra-individual mode distributions. This localized analysis was used to quantify the percentage of participants exhibiting multiple distinct morphologies across different anatomical sites.

### 2.8 Statistical Analysis

Statistical analyses were conducted using Python (v3.12) utilizing the statsmodels and scipy.stats libraries. All analyses were performed exclusively on the Training Set (*n* = 219) to avoid data leakage before independent model validation.

Baseline demographic and clinical characteristics were compared between the non-hypertensive and hypertensive cohorts using Welch’s *t*-test for numerical variables (including age, weight, and Monk Skin tone) and Pearson’s Chi-square test for categorical variables (specifically biological sex). Welch’s test variants were specifically selected to ensure statistical robustness against unequal variances and distribution deviations within this dataset.

To evaluate the relationship between physiological habitus and sensor morphology, systolic peak amplitude (*A*_sp_) was compared across body mass index (BMI) categories using Welch’s one-way ANOVA followed by a Games–Howell post-hoc pairwise test. For the purpose of statistical testing, the Underweight and Healthy weight categories were merged into a single “Healthy/Underweight” group (BMI *<* 25 kg/m^2^). This adjustment was necessary to ensure sufficient sample sizes for robust variance estimation, as the Underweight cohort alone (*n ≤* 6) lacked the required power for independent analysis. A two-way ANOVA was subsequently applied to evaluate the main and interaction effects of BMI category and BP status on *A*_sp_.

For timing-based Pulse Wave Analysis (PWA), we investigated three features mapping the systolic phase to capture surrogate markers of arterial stiffness: time from onset to systolic peak (*T*_sp_), time from onset to *u*-point (*T*_u_), and time from onset to *b*-point (*T*_b_) - normalizing each by pulse duration (*T*_pi_) to remove potential confounding effects from heart rate. To segment distinct physiological profiles within this timing feature space, Gaussian Mixture Models (GMM) were deployed. The number of mixture components was fixed at two based on the clear, empirical bimodality observed in the feature distributions. Finally, the prevalence of the steepest morphological modes across different BMI and BP groups was evaluated using a Generalized Linear Model (GLM) with a binomial distribution and a logit link function, without adjustment for additional demographic covariates such as age or biological sex; significance was assessed via the Wald chi-squared test. Statistical significance was defined as *p <* 0.05 unless otherwise specified.

### 2.9 Classification Framework and Feature Selection

To evaluate the predictive value of sensor-derived features for hypertension classification, we developed three low-complexity machine learning models: Logistic Regression, Random Forest, and Support Vector Machines (SVM) with a radial basis function (RBF) kernel. Model performance was evaluated using stratified 5-fold cross-validation restricted entirely to the Training Set.

Classification was executed across two distinct phases: a demographic-only baseline (age, weight, height, heart rate, and biological sex) and a sensor-augmented framework to calculate the change in the Area Under the Receiver Operating Characteristic curve (ΔAUC). The sensor feature space comprising 83 standard PWA features was derived from the representative signal templates using the pyPPG software library [14] and morphological mode prevalence derived from the unsupervised archetype pipeline.

High-dimensional feature pruning was performed using *L*_1_ regularization (Lasso) for Logistic Regression, tree-based impurity importance ranking combined with an incremental forward-selection wrapper for Random Forest, and permutation importance for the RBF-SVM. The independent hold-out test set remained completely isolated and untouched to maintain strict methodological generalizability for future work.

## 3 Results

### 3.1 Participant Characteristics and Clinical Baseline

Baseline demographic and clinical characteristics for the study cohort are summarized in Table 1. The hypertensive cohort was significantly older (52.0 *±* 10.6 vs. 47.3 *±* 12.5 years, *p* = 0.001) and exhibited a higher mean BMI (29.4 *±* 6.5 vs. 25.9 *±* 5.1 kg/m^2^, *p <* 0.001) compared to the non-hypertensive cohort. Evaluation of skin characteristics revealed significantly lower Colorimeter ITA values in the HTN_True group (55.2 *±* 8.2 vs. 58.7 *±* 7.7 *^◦^*, *p* = 0.004), indicating a higher prevalence of darker skin tones, though subjective Monk Skin Tone classifications did not differ significantly between groups (4.1 *±* 1.8 vs. 3.8 *±* 1.7, *p* = 0.209).

**Table 1:** Baseline Demographics and Clinical Characteristics of the Study Cohort.

| Category | Feature | Non-Hypertension | Hypertension | p-value |
| --- | --- | --- | --- | --- |
|  |  | n = 131 | n = 130 |  |
| Demographics | Age (years) | 47.3 (12.5) | 52.0 (10.6) | 0.001* |
|  | Height (cm) | 171.4 (9.9) | 170.6 (9.8) | 0.531 |
|  | Weight (kg) | 76.7 (19.1) | 85.9 (21.4) | < 0.001* |
|  | BMI (kg/m<sup>2</sup>) | 25.9 (5.1) | 29.4 (6.5) | < 0.001* |
|  | Colorimeter ITA (°) | 23.4 (21.4) | 14.9 (22.8) | 0.004* |
|  | Monk Skin Tone | 3.8 (1.7) | 4.1 (1.8) | 0.209 |
|  | Skin Hydration (a.u.) | 24.4 (13.6) | 24.9 (14.1) | 0.775 |
|  | Female, n (%) | 51 (38.9%) | 46 (35.4%) | 0.52 |
|  | Male, n (%) | 80 (61.1%) | 83 (63.8%) |  |
|  | Not-Specified, n (%) | 0 (0%) | 1 (0.8%) | NA |
| Blood Pressure and Heart Rate | Systolic BP (mmHg) | 113.0 (9.8) | 143.7 (14.3) | < 0.001* |
|  | Diastolic BP (mmHg) | 68.7 (7.3) | 87.1 (8.6) | < 0.001* |
|  | Heart Rate (bpm) | 73.6 (12.6) | 77.4 (12.5) | 0.015* |

Continuous variables are expressed as mean (standard deviation) and categorical variables as count (percentage). Statistical significance between the non-hypertensive and hypertensive cohorts was evaluated using Welch’s t-test for continuous variables and Pearson’s chi-square test for categorical variables. Asterisks (\*) denote statistical significance ( $p < 0.05$ ). a.u. = arbitrary units.

Per the stratification criteria, both mean systolic (143.7 *±* 14.3 vs. 113.0 *±* 9.8 mmHg, *p <* 0.001) and diastolic (87.1 *±* 8.6 vs. 68.7 *±* 7.3 mmHg, *p <* 0.001) BP were significantly elevated in the hypertensive cohort. Mean heart rate was also significantly elevated in the hypertensive participants (77.4 *±* 12.5 vs. 73.6 *±* 12.6 bpm, *p* = 0.015).

Cross-correlation analysis of these baseline demographic and clinical variables (Supplementary Fig. S1) demonstrated a strong relationship between BMI and total body weight (*r* = 0.89). As anticipated, extreme collinearity was observed between systolic and diastolic pressure (*r* = 0.81). Additionally, a moderate negative correlation was observed between objective Colorimeter ITA values and subjective Monk Skin Tone classifications (*r* = *−*0.61).

### 3.2 Comparative Analysis of IPG and PPG Waveform Morphology

To evaluate the morphological variations captured by each sensing modality, time-/amplitude-normalized and ensemble-averaged cardiac cycle templates were compared across cohorts, anatomical sites, and BMI strata. Figure 2 illustrates the cohort-wide mean waveforms (*±*1 standard deviation) grouped by BP status. A striking divergence in spatial distribution was observed between the two sensing modalities. PPG waveforms exhibited a highly conserved, uniform unimodal morphology across all three anatomical locations, displaying minimal variation between the non-hypertensive and hypertensive cohorts. In contrast, IPG signals demonstrated pronounced, site-specific morphological complexity. At the ventral forearm reference site, the IPG waveform resembled a central arterial pressure wave, characterized by a sharp systolic upstroke followed by a distinct secondary diastolic peak. Moving distally to the ventral wrist, the IPG waveform retained this rapid upstroke but introduced high-frequency dicrotic reflections (inflection ripples) along the downward slope. At the dorsal wrist, the IPG signal represented a hybrid profile, tracking as a spatial superposition of the ventral arterial components and the localized tissue volume changes characteristic of the co-located PPG signal.

**Figure 2:**
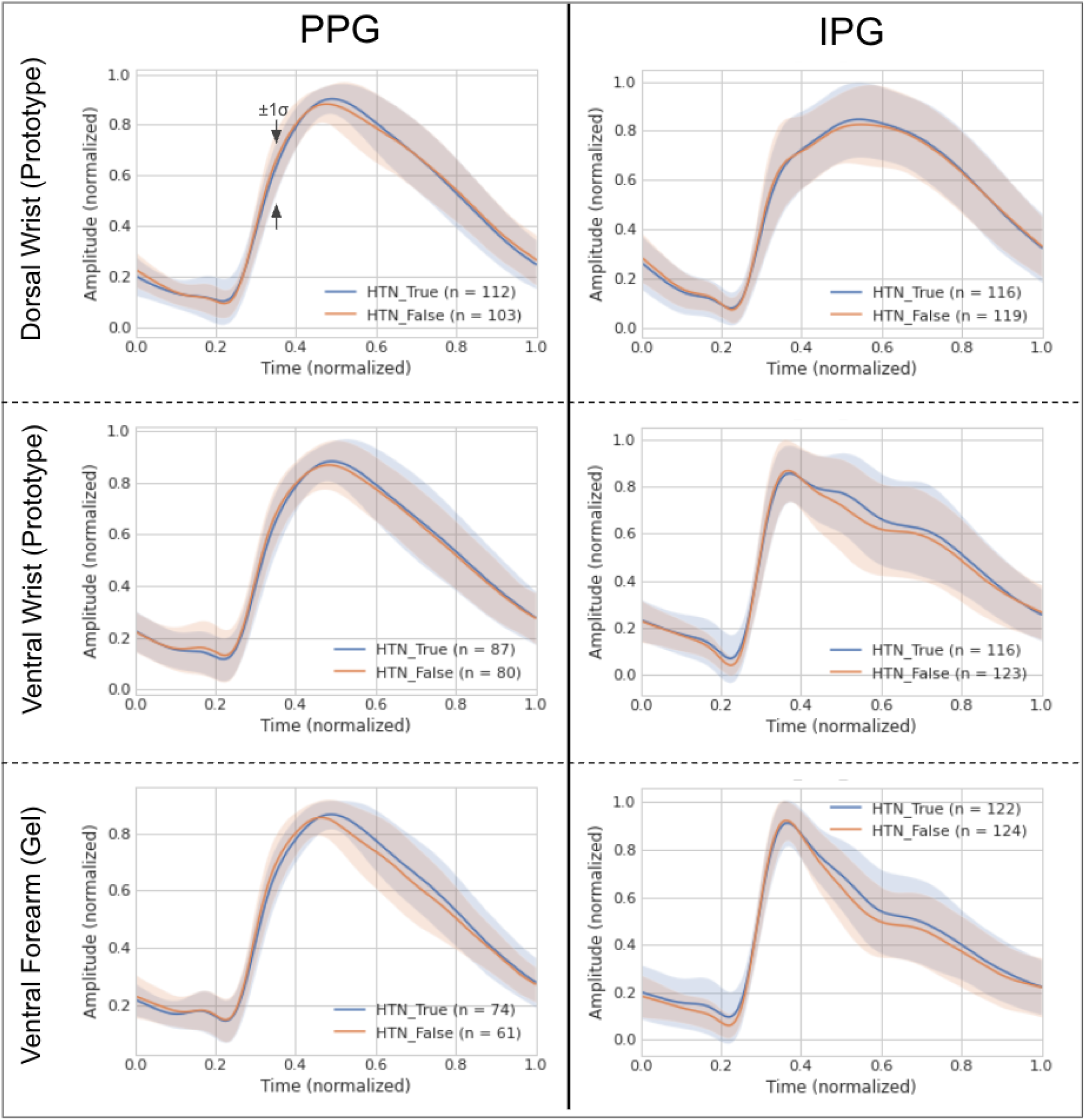
Comparative morphological analysis of PPG versus IPG across anatomical sites. Ensemble-averaged, amplitude-normalized cardiac cycles (mean ± 1 standard deviation) stratified by hypertensive (HTN_True, blue) and non-hypertensive (HTN_False, orange) cohorts. Analysis is performed for both PPG (left) and IPG (right) across the dorsal wrist prototype (top), ventral wrist prototype with saline (middle), and ventral forearm gel electrodes (bottom). While the optical PPG signal (left column) exhibits a highly conserved, uniform morphology regardless of measurement site or clinical status, the IPG signal (right column) demonstrates pronounced spatial heterogeneity. IPG resolves complex, site-specific hemodynamic features that remain invisible to PPG, particularly at the ventral forearm reference site.

These modality-specific variations were mirrored in the first and second time derivatives (Supplementary Figs. S2 and S3). Despite distinct mean trajectory shifts between BP cohorts, a substantial statistical overlap was present across all subplots, demonstrating a likely challenge for hypertension classification. Lastly, there were notably more high-quality beats with IPG than PPG for comparing the waveform morphologies (Fig. 2) - e.g., for the ventral forearm (gel) measurements, IPG had *>* 1.5*×* participants with high quality beats in the HTN_True cohort compared to PPG, and *>* 2.0*×* for the HTN_False cohort.

Subgroup analysis stratified by CDC-standard BMI categories further exposed the sensitivity of IPG to physiological habitus compared to the homogeneous response of PPG (Fig. 3; Note: Due to a low sample size in the underweight cohort (*n ≤* 6), its wider variance should be interpreted with caution). While the PPG waveforms retained an identical unimodal structure across all weight classes, the IPG waveforms revealed a clear physiological gradient. At the ventral forearm, the primary systolic upstroke became progressively attenuated and “dull” as body habitus advanced from healthy weight to obesity. Furthermore, while a distinct diastolic peak was clearly resolved in healthy-weight and underweight individuals, it transitioned into an elevated “tidal wave” profile in the mid-diastolic phase among overweight and obese participants. This tidal wave component progressively elevated and smoothed out as the sensor placement moved from the ventral forearm to the ventral and dorsal wrist, reflecting a mass-dependent damping of peripheral hemodynamics. IPG waveforms showed clear morphological stratification across BMI categories (healthy/underweight, overweight, and obese), whereas PPG morphology remained relatively homogeneous across the same categories.

**Figure 3:**
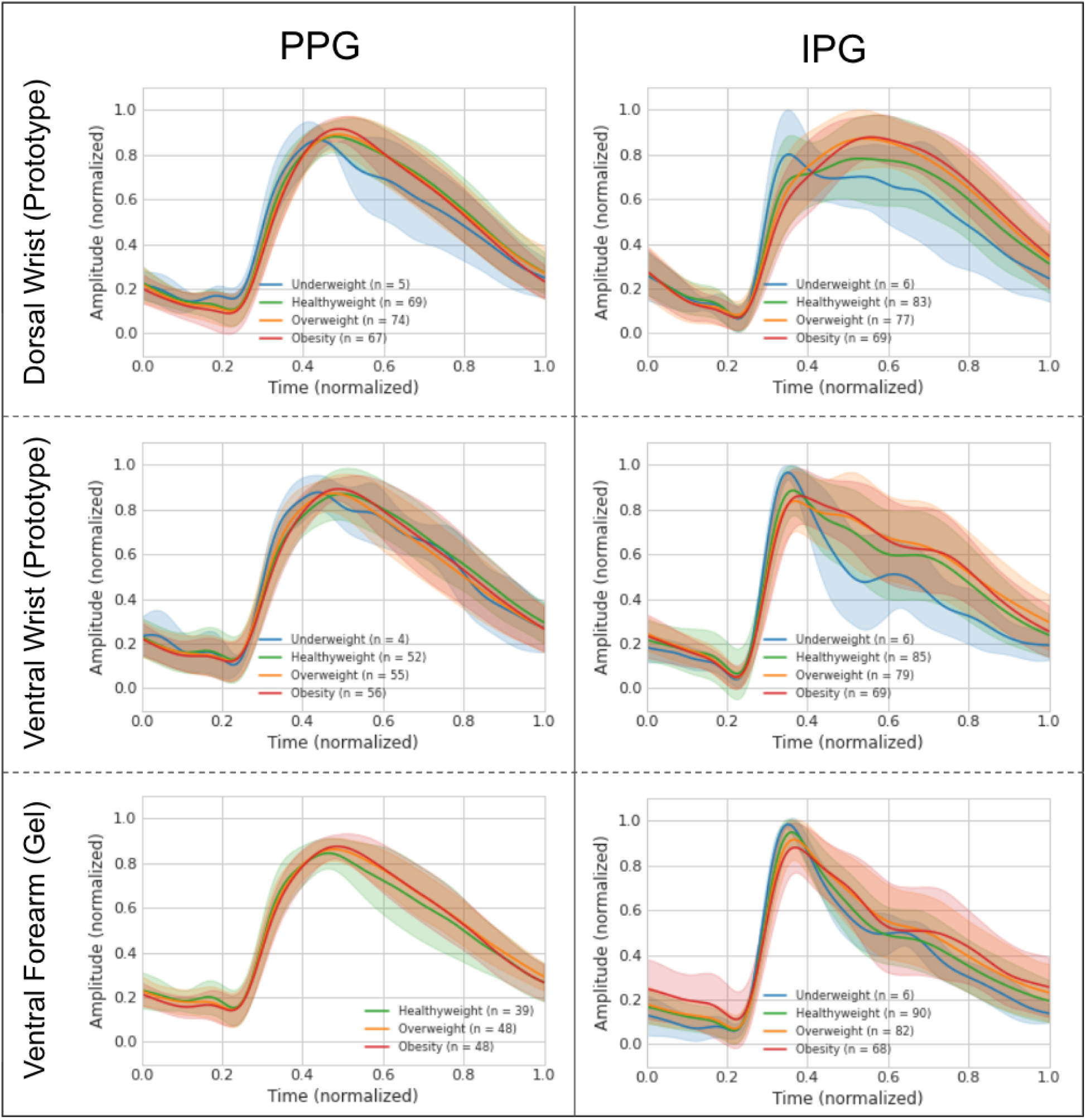
Influence of physiological habitus (BMI) on PPG and IPG waveform morphology. Ensemble-averaged waveforms (mean ± 1 standard deviation) stratified by CDC-standard BMI categories: underweight (blue), healthy weight (green), overweight (orange), and obesity (red). Analysis is performed for both PPG (left) and IPG (right) across the dorsal wrist prototype (top), ventral wrist prototype with saline (middle), and ventral forearm gel electrodes (bottom). PPG waveforms (left column) remain fundamentally homogeneous across all weight classes. In contrast, IPG (right column) reveals a distinct physiological gradient. This sensitivity is most evident at the ventral forearm, where increasing BMI systematically attenuates the primary systolic upstroke and elevates the mid-diastolic profile, quantifying the damping effect of adipose tissue on deep-tissue sensing.

### 3.3 Modality Sensitivity to BMI and Blood Pressure

To investigate the impact of physical habitus on signal magnitude, we evaluated the systolic peak amplitude (*A*_sp_) across BMI and BP categories (Fig. 4). One-way ANOVA revealed that *A*_sp_ was significantly modulated by BMI category for both sensors at the dorsal wrist (PPG: p = 0.010; IPG: p = 0.008) and most notably for IPG at the ventral forearm reference site (p < 0.001; Supplementary Table 1). Games-Howell post-hoc testing indicated that these differences were primarily driven by the variance between the Healthy/Underweight and Obese groups (Supplementary Table 2).

**Figure 4:**
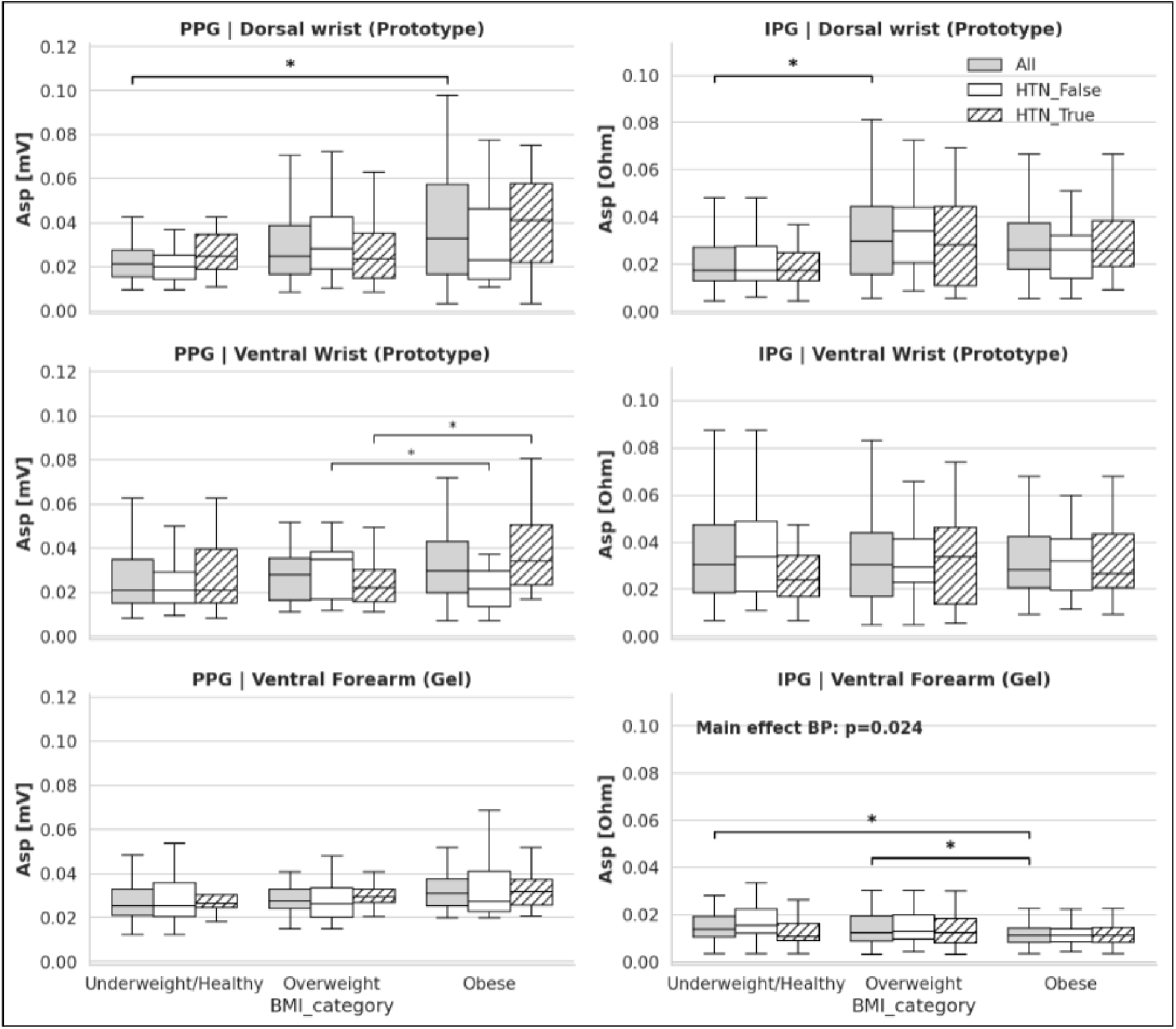
Sensitivity of systolic peak amplitude (*A*_sp_) to body mass index (BMI) and blood pressure (BP) status. Boxplots represent the distribution of pulsatile amplitudes for PPG (left column, mV) and IPG (right column, Ohm) stratified by pooled BMI categories. Distributions are further partitioned into the aggregate cohort (All, grey), non- hypertensive participants (HTN_False, white), and hypertensive participants (HTN_True, hatched) across the dorsal wrist prototype (top), ventral wrist prototype with saline (middle), and ventral forearm gel electrodes (bottom). Notably, IPG at the ventral forearm (bottom right) is the only sensor-site configuration to exhibit a significant main effect for BP status (p = 0.024). Asterisks (*) note statistically significant pairwise differences (p < 0.05) between BMI categories.

When introducing BP status as a second factor in a two-way ANOVA, we observed that the IPG ventral forearm was the only configuration to exhibit a significant main effect of BP status on amplitude (p = 0.024; Fig. 4). No other sensor-site combination demonstrated this direct sensitivity to hypertension status. While PPG at the ventral wrist did not show a main effect of BP, it exhibited a significant interaction between BP and BMI group (p = 0.002). Post-hoc analysis of this interaction showed that for both non-hypertensive participants and hypertensive participants, *A*_sp_ differed between Overweight and Obese cohorts (p = 0.020 and p = 0.009 respectively; Supplementary Tables 3 and 4).

### 3.4 Pulse Wave Timing Features Distributions and Latent Profiles

Analysis of normalized timing features (*T*_sp_*/T*_pi_, *T*_u_*/T*_pi_, and *T*_b_*/T*_pi_) revealed distinct distributional bimodalities within the feature space (Fig. 5). *T*_b_*/T*_pi_ was omitted from visualization due to its extremely high correlation with *T*_u_*/T*_pi_. GMM applied to these timing features segmented participants into two distinct latent morphological profiles: an A-line-like morphology (Profile 0) and a tidal-dominant morphology (Profile 1).

**Figure 5:**
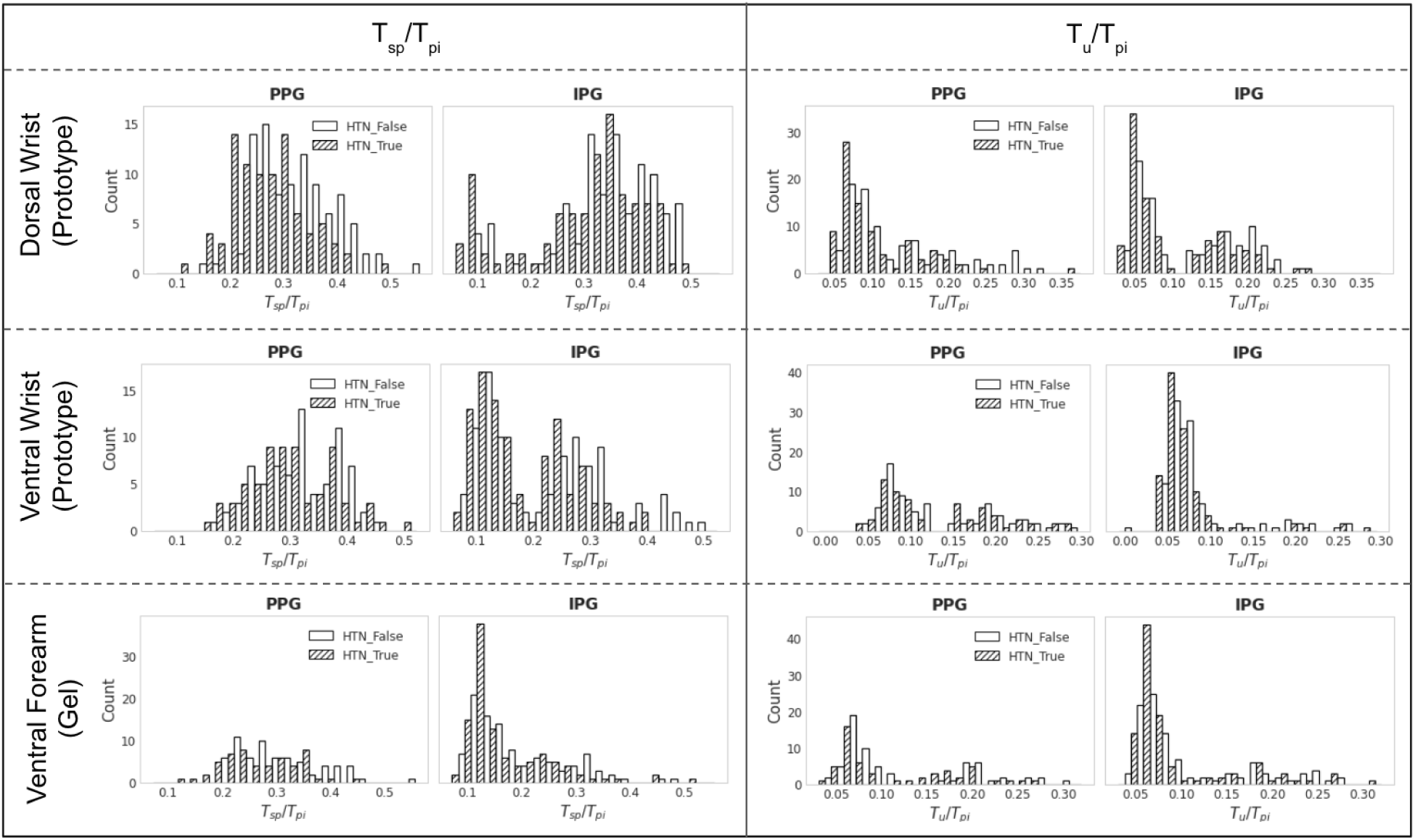
Population-wide distribution of normalized pulse wave timing features. Histograms illustrate the frequency distribution of the normalized time from pulse onset to systolic peak (*T*_sp_*/T*_pi_; left) and time to max slope (*T*_u_*/T*_pi_; right) across PPG and IPG modalities. Data are stratified by hypertensive (HTN_True, hatched bars) and non-hypertensive (HTN_False, white bars) across the three anatomical measurement sites - dorsal wrist prototype (top), ventral wrist prototype with saline (middle), and ventral forearm gel electrodes (bottom). While PPG feature distributions remain largely homogeneous, IPG feature spaces reveal a distinct structural bimodality.

At the ventral forearm reference site, IPG Profile 0 closely mimicked an A-line waveform, characterized by a rapid upstroke and distinct dicrotic resolution, whereas IPG Profile 1 showed a more “damped” tidal-wave appearance (Fig. 6). In contrast, co-located PPG waveforms remained visually indistinguishable between Profile 0 and Profile 1. Spatially, the IPG Profile 0 retained its A-line-like characteristics at the ventral wrist, while Profile 1 introduced inflection ripples along the diastolic slope. At the dorsal wrist, IPG Profile 0 showed potential waveform augmentation, and IPG Profile 1 converged toward a unimodal morphology similar to the PPG signal.

**Figure 6:**
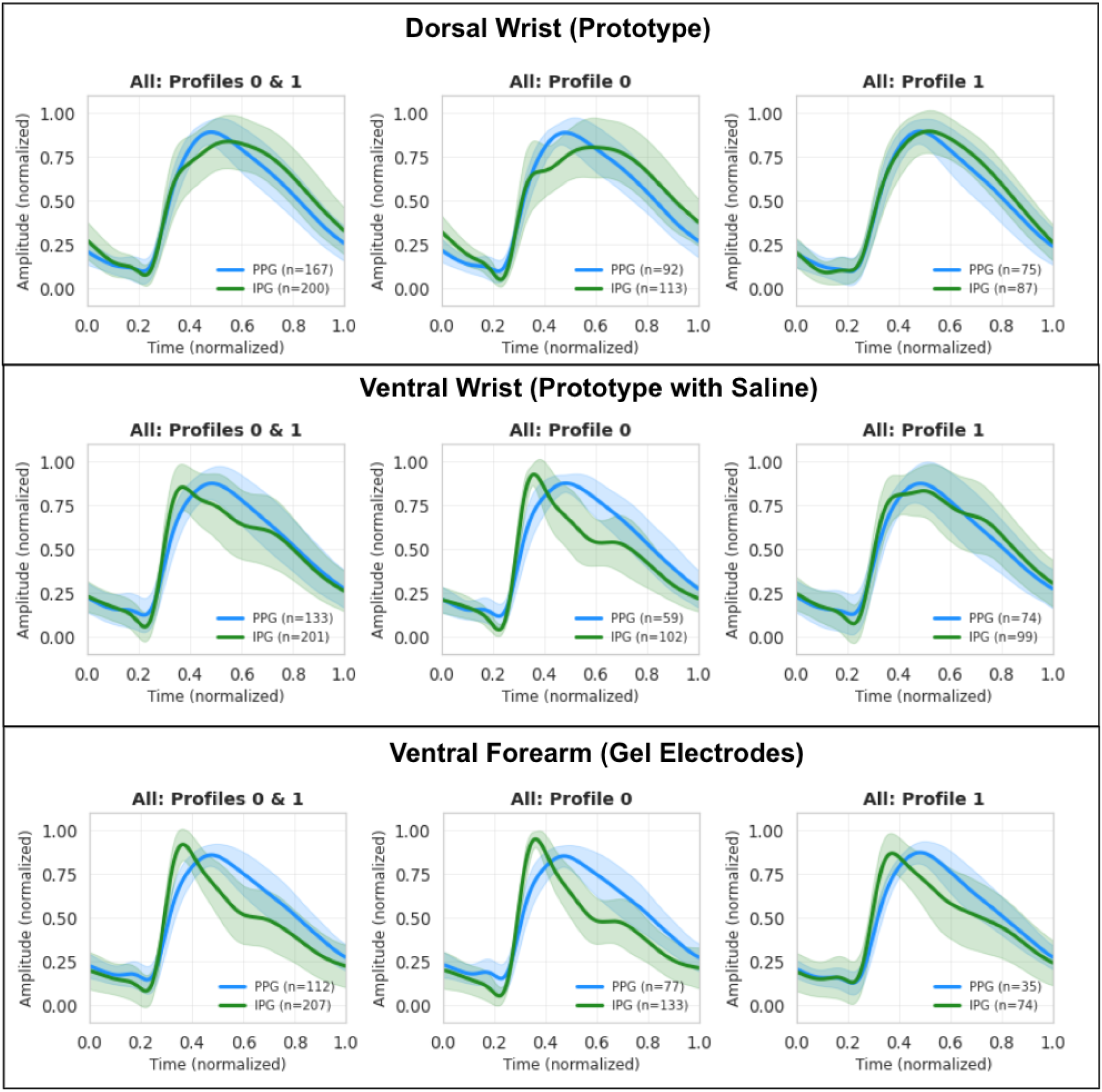
Stratification of aggregated waveforms into latent morphological profiles via Gaussian Mixture Models (GMM). Ensemble-averaged waveforms (mean *±*1 standard deviation) for PPG (blue) and IPG (green) were segmented across three anatomical locations: dorsal wrist prototype (top), ventral wrist prototype with saline (middle), and ventral forearm gel electrodes (bottom) into two distinct morphological modes using a GMM framework trained on pulse wave timing features (*T*_sp_*/T*_pi_, *T*_u_*/T*_pi_, and *T*_b_*/T*_pi_). The first column illustrates the pooled, unstratified dataset for each modality. The subsequent columns isolate the discovered latent profiles. Notably, IPG clearly reveals an A-line-like morphology (Profile 0) distinct from a damped, tidal-dominant morphology (Profile 1). In stark contrast, co-located PPG waveforms remain visually indistinguishable between the two profiles. (Note: Profile assignment was restricted to participants exhibiting a GMM confidence level *>* 0.85).

Stratification of these latent profiles by BP cohort revealed site-specific pathological signatures in Profile 0 (Supplementary Fig. S4). At the dorsal wrist, hypertensive individuals within Profile 0 exhibited an “augmented squareness” - a sustained plateau in the mid-diastolic region - contrasted with the more rapid decay observed in non-hypertensive participants. At the ventral wrist, the hypertension cohort demonstrated a significantly more pronounced tidal wave component during the mid-to-late diastolic phase. These cohort-specific divergences were absent in Profile 1 and at the ventral forearm reference site.

### 3.5 Unsupervised Archetype Discovery and Beat Heterogeneity

Following the observation of latent morphological variations in the aggregated templates, we sought to quantify the intra-individual, beat-to-beat heterogeneity using the unsupervised archetype discovery pipeline. To ensure statistical representativeness while accommodating the majority of physiological variance, a subset of 100 cardiac cycles was randomly sampled per participant for each sensor modality and anatomical site (Supplementary Fig. S5).

Using a Silhouette threshold of 0.25 to promote cluster separation, global analysis revealed that IPG consistently resolved more complex morphological modes than PPG (Fig. 7). At the dorsal wrist, IPG clustered into three distinct archetypes - ranging from sharp, arterial-like peaks to wider, elevated mid-diastolic profiles. Conversely, the co-located PPG signals converged into two highly conserved unimodal shapes. The prevalence of these emergent IPG archetypes was significantly influenced by BMI. For IPG recorded at the ventral forearm, the prevalence of the steeper morphological variant (Mode 1) was significantly lower in the obese group (*p* = 0.035; Fig. 8, Supplementary Table 5). Some PCA-derived archetypes appeared morphologically similar to the profiles calculated from cross-correlation (Supplementary Fig. S6); however, there were also clear modes that were distinct and cases where these modes were very morphologically different to the cross-correlation profiles (almost appearing as a deconstructed signal) such as for IPG dorsal wrist prototype.

**Figure 7:**
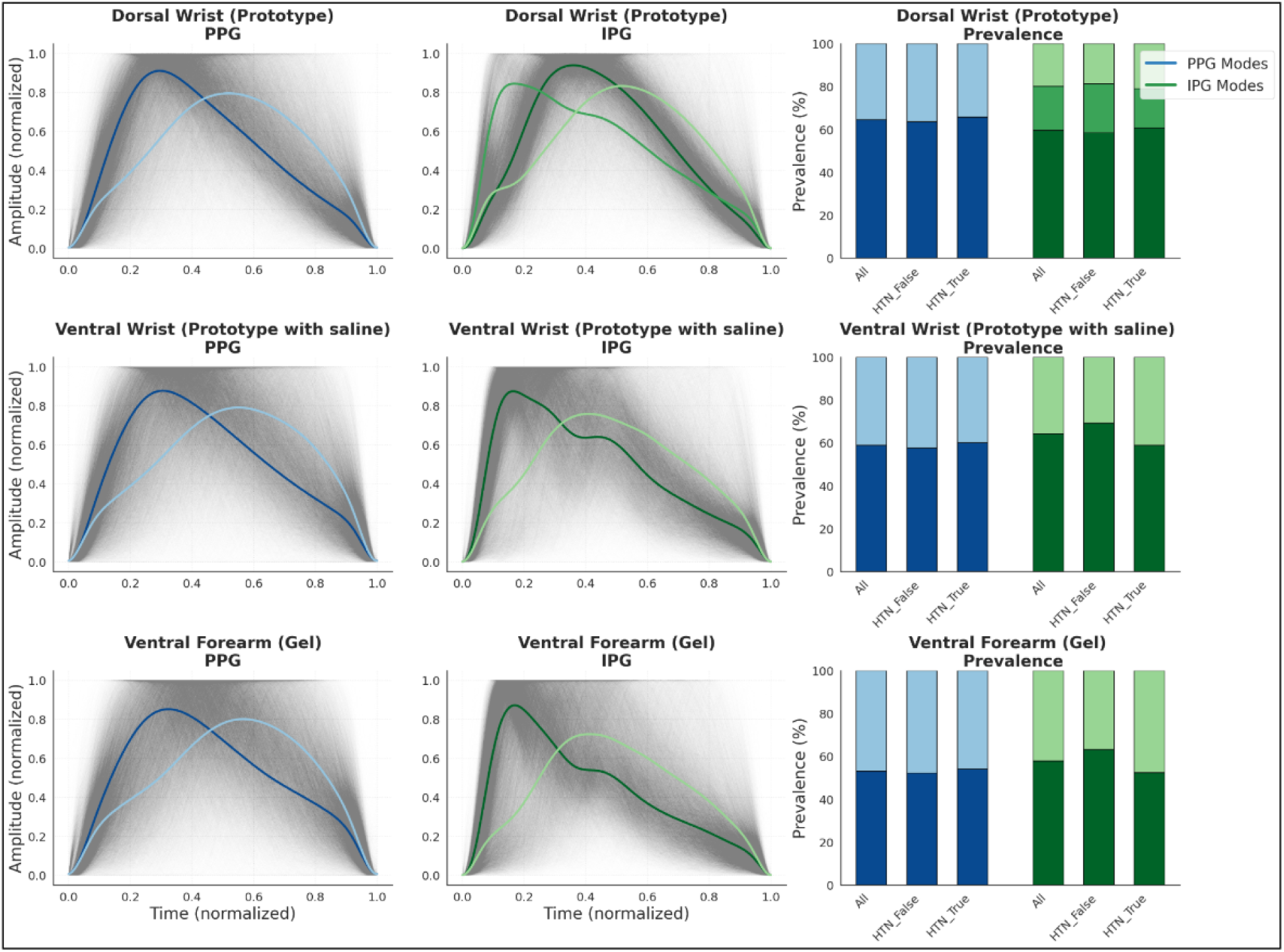
Unsupervised global archetype discovery and morphological mode prevalence. Representative morphological archetypes discovered via principal component analysis (PCA) and K-means clustering for PPG (left columns, blue) and IPG (middle column, green) across three anatomical sites: dorsal wrist prototype (top), ventral wrist prototype with saline (middle), and ventral forearm gel electrodes (bottom). The right column displays the relative prevalence of these discovered archetypes, stratified by hypertension status. IPG consistently resolves a higher degree of morphological complexity - identifying up to three distinct structural archetypes at the dorsal wrist - whereas PPG signals uniformly converge into two nearly identical, unimodal shapes.

**Figure 8:**
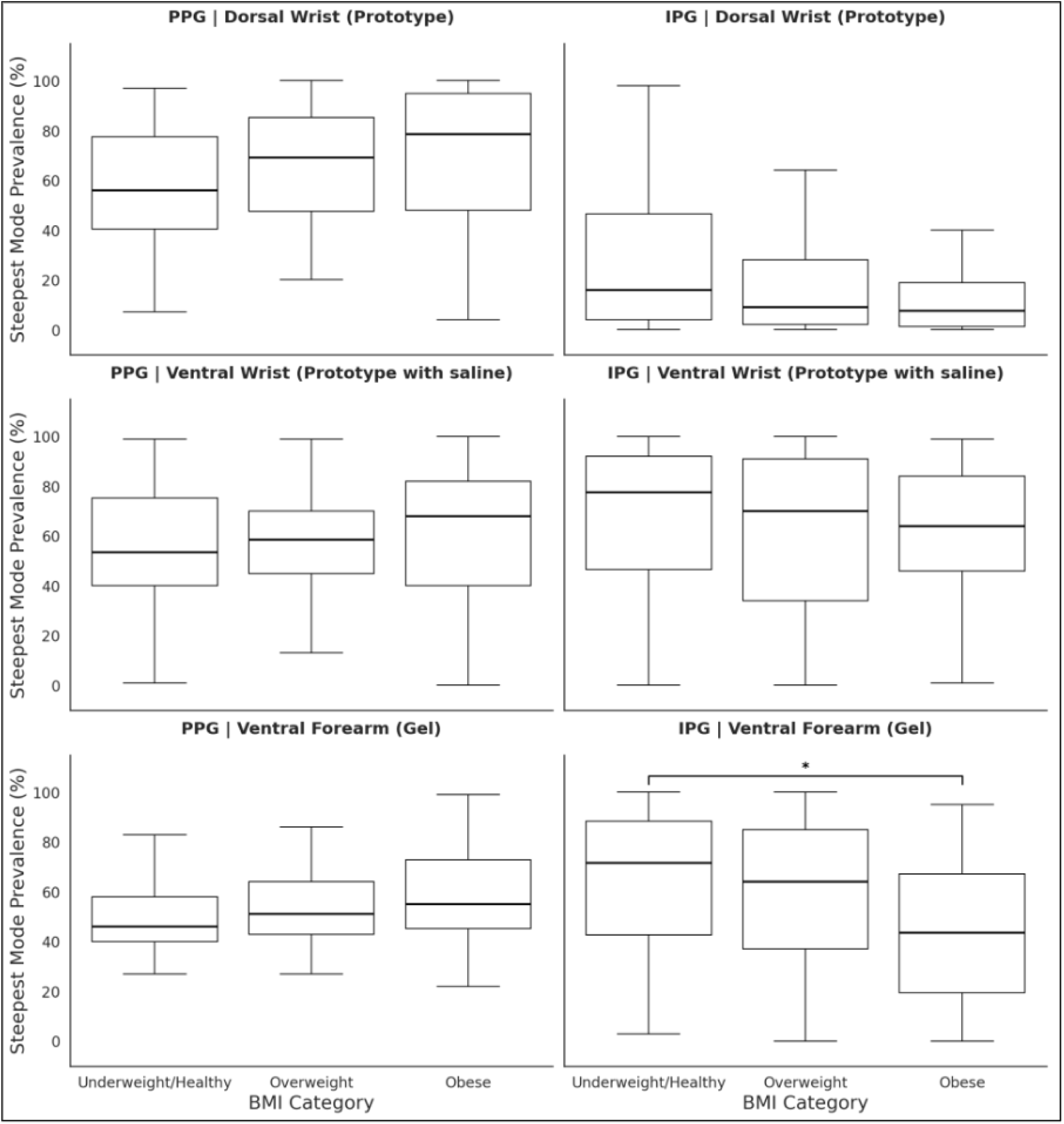
Impact of physiological habitus (BMI) on the prevalence of steep pulsatile morphological modes. Boxplots illustrate the prevalence (%) of the steepest identified pulse mode (e.g., Mode 1) across Underweight/Healthy, Over-weight, and Obese BMI categories. Data are partitioned by modality into PPG (left column) and IPG (right column) across three anatomical sites: dorsal wrist prototype (top), ventral wrist prototype with saline (middle), and ventral forearm gel electrodes (bottom). Notably, the prevalence of the steepest IPG mode at the ventral forearm is significantly attenuated in the obese cohort compared to the healthy/underweight cohort (* p < 0.05), quantifying the damping effect of subcutaneous adipose tissue on IPG signals.

To assess the robustness of these discovered archetypes, an individual-level clustering threshold sweep was performed (Fig. 9). As the Silhouette threshold - a hyperparameter for cluster cohesion - was increased, the percentage of participants exhibiting multiple modes (*k >* 1) decreased more rapidly for PPG than for IPG across all anatomical sites. At a threshold of 0.4 - selected because it captured a stable 80/20 distribution in ventral wrist PPG - IPG continued to resolve multiple distinct morphologies (e.g., dual systolic wave profiles) that were obscured in the PPG signal space. Across the threshold sweep, the rate of multi-mode attenuation in the PPG signals exhibited a spatial dependence, dropping most rapidly at the ventral forearm, followed by the ventral wrist prototype, and lastly the dorsal wrist prototype. Conversely, the IPG multi-mode distributions remained robustly distributed and less sensitive to spatial site transitions.

**Figure 9:**
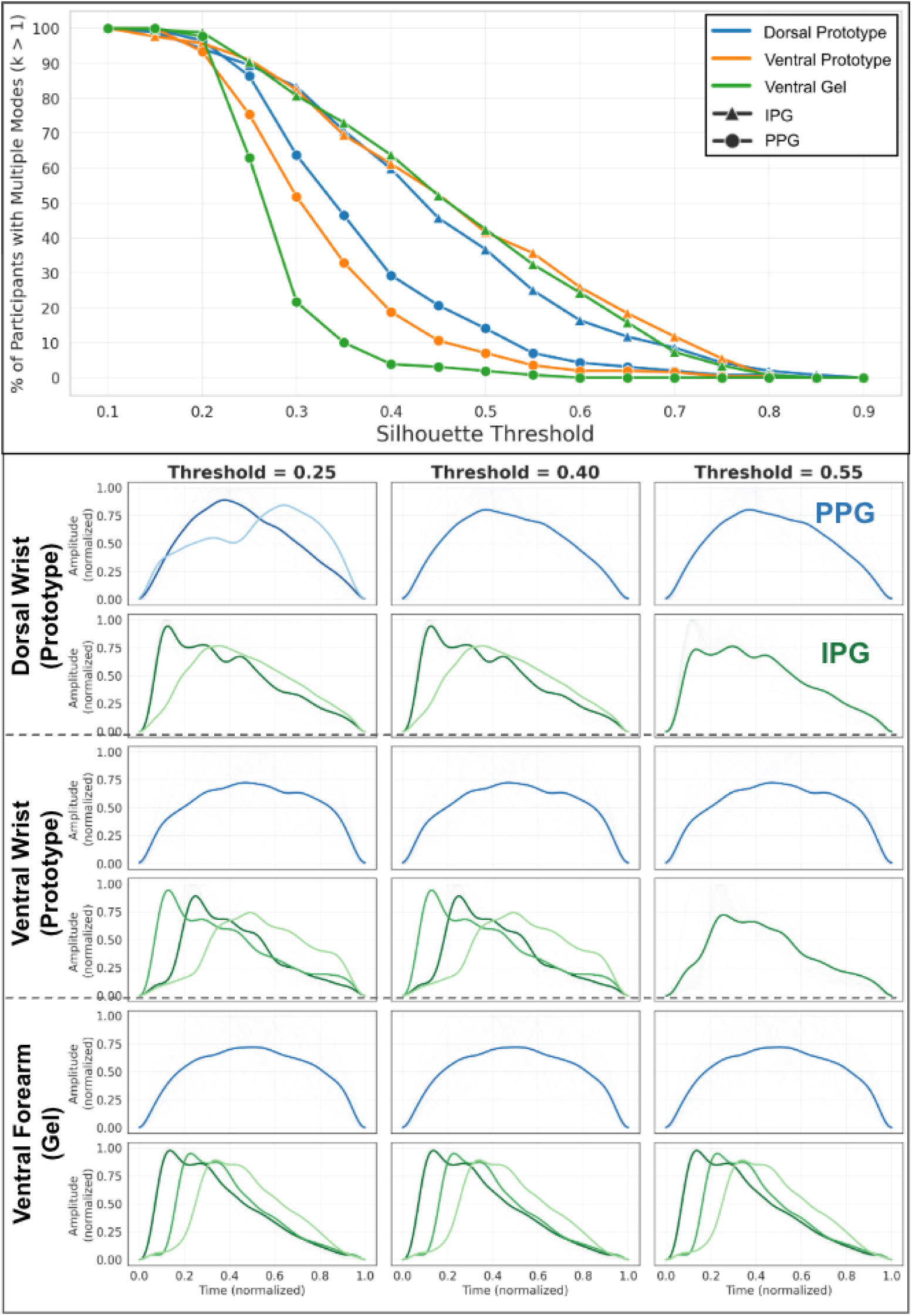
Sensitivity and robustness of unsupervised morphological mode discovery. (Top) A threshold sweep illustrating the percentage of participants exhibiting multiple distinct waveform modes (k > 1) as a function of the Silhouette threshold. Across all measurement sites, PPG signals (circles) rapidly converge to a single mode, whereas IPG signals (triangles) demonstrate robust intra-individual heterogeneity that persists at higher clustering thresholds. (Bottom) Representative intra-individual waveform archetypes from a single participant visualized across increasing Silhouette thresholds (0.25, 0.4, 0.55). At a strict threshold of 0.40, PPG (blue) collapses into a single morphological shape, while IPG (green) resolves and maintains multiple distinct hemodynamic modes (e.g., dual systolic wave profiles).

This fundamental difference in signal richness is most clearly observed in the distribution of individual mode counts at the 0.4 threshold (Supplementary Fig. S7). Across all anatomical sites, PPG was heavily skewed toward a singular morphological mode (*k* = 1) for the vast majority of participants. In contrast, IPG demonstrated a higher degree of intra-individual heterogeneity, with the majority of participants resolving two or more distinct morphologies within a single recording. This trend was particularly pronounced at the ventral forearm (gel), where PPG converged almost entirely into a single mode, while IPG maintained a broad distribution of participants with two or three unique beat archetypes.

### 3.6 Hypertension Classification Performance

The incremental predictive value of the engineered pulse wave features was evaluated across a series of low-complexity classifiers, with the SVM yielding the highest overall cross-validated performance. Classification results across data modalities, anatomical sites, and feature extraction frameworks are illustrated via Receiving Operating Characteristic curves in Fig. 10. A comprehensive summary of these classification results is provided in Table 2, which quantifies the peak AUC scores and highlights the specific pulse wave features that provided incremental predictive value over the demographic baseline.

**Figure 10:**
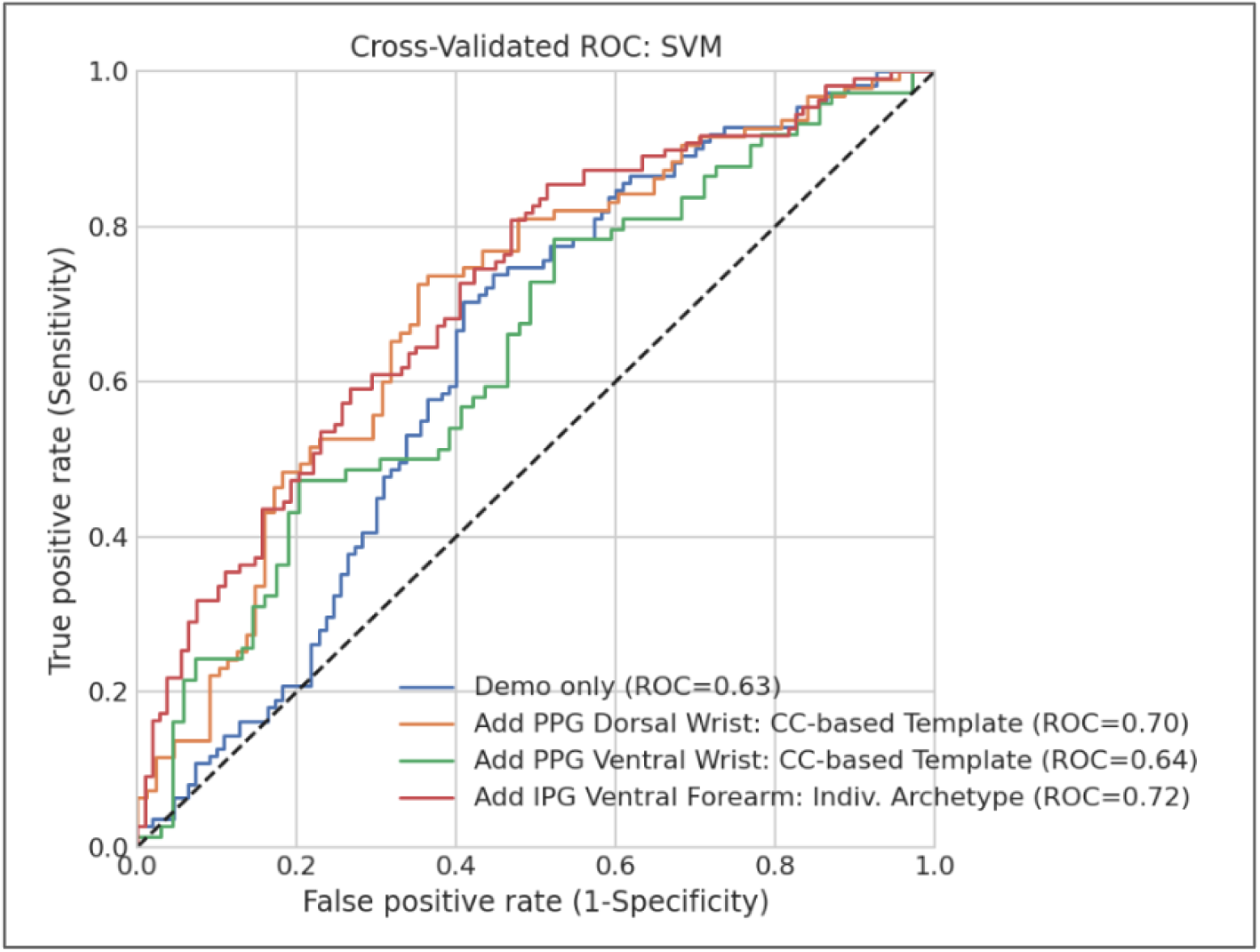
Hypertension classification performance demonstrating the incremental predictive value of sensor-derived features. Cross-validated Receiver Operating Characteristic (ROC) curves for Support Vector Machine (SVM) models evaluated on the Training Set. The baseline model utilizing only demographic features (age, weight, height, heart rate, and biological sex) achieved an AUC of 0.63 (blue line). Appending traditional cross-correlation-based PPG features from the dorsal wrist yielded a modest improvement (AUC = 0.70, orange line). Integrating individual IPG morphological archetypes extracted from the ventral forearm reference site (AUC = 0.72, red line) shows the distinct pathological signatures encoded in deep-tissue bioimpedance.

**Table 2:**
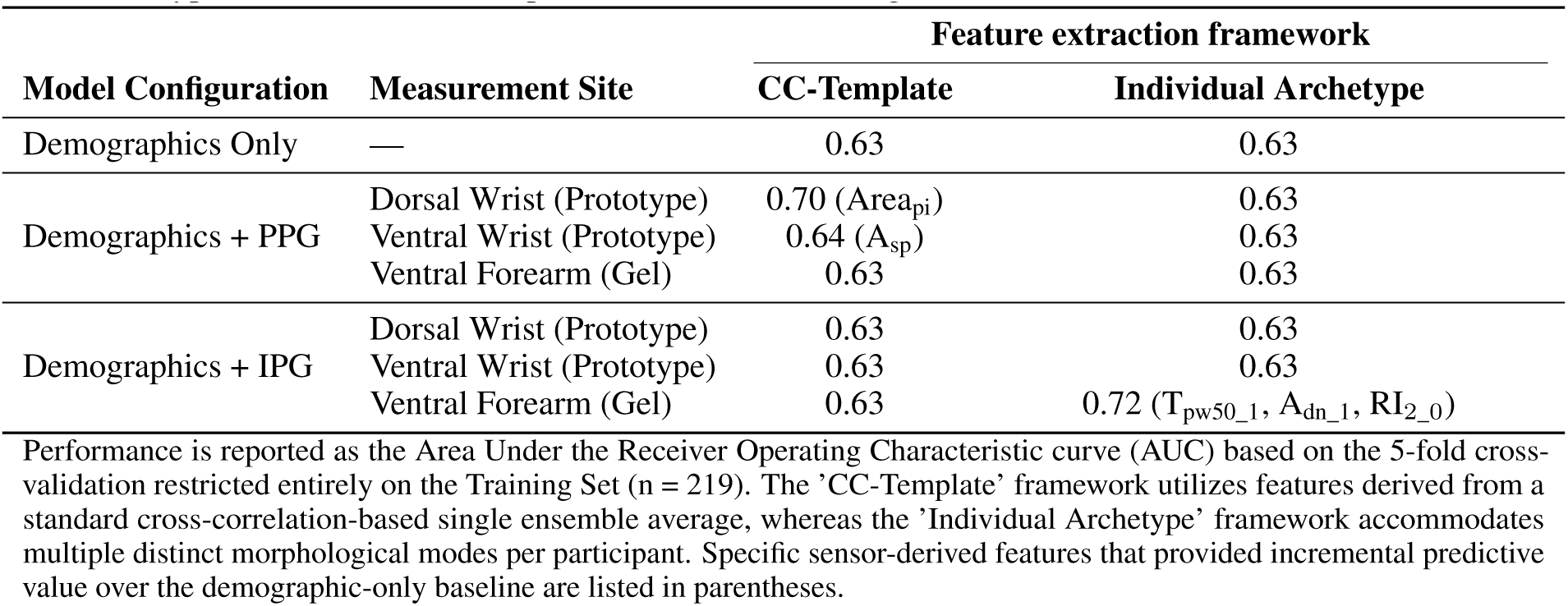
Hypertension classification performance across sensing modalities and feature extraction frameworks.

A baseline model restricted exclusively to demographic variables (weight, age, height, heart rate, and biological sex) achieved an AUC of 0.63. The integration of sensor-derived features provided distinct, modality-dependent shifts in the ROC space, pulling the performance curves towards the upper-left quadrant. Notably, the extraction frameworks behaved inversely between the two modalities:

- PPG: Traditional PWA metrics derived from standard cross-correlation (CC) waveform templates yield modest improvements over the demographic baseline, with the dorsal wrist prototype achieving the highest PPG-based performance (AUC = 0.70), driven primarily by the pulse interval area (Area_pi_). Conversely, clustering-based archetype extraction yielded no predictive uplift for the PPG modality.
- IPG: PWA features derived from CC waveform templates failed to improve upon the demographic baseline. However, executing the unsupervised pipeline at the individual level unlocked the highest classification performance in the study. Appending individual archetypes from the ventral forearm (gel) configuration achieved an AUC of 0.72. This performance peak was driven by fine-grained morphological landmarks

that were uniquely resolvable through individual-based clustering, specifically, the 50% pulse width of the first mode (*T*_pw50_1_), the diastolic notch amplitude (*A*_dn_1_), and the second reflection index (RI_2_0_). For this individual pipeline, a maximum allocation of two modes (*k ≤* 2) per participant was enforced, a constraint supported by the empirical mode distributions shown in Supplementary Fig. S7, where the vast majority of participants across both BP cohorts exhibited two or fewer dominant morphologies.

In contrast to the success of individual-level tracking, features derived from global-level population archetypes failed to provide any predictive uplift over the baseline demographic model across all sites and modalities. To maintain model parsimony and optimize reporting clarity, this uninformative data was excluded from the final comparative matrix.

While the absolute performance gains achieved by integrating sensor metrics remain modest, all reported classification metrics represent internal 5-fold cross-validation restricted entirely to the Training Set. Because the cross-validated performance delta did not reach the high threshold required to justify model freezing, the independent hold-out Test Set (*n* = 42) remains entirely intact, leaving an uncompromised benchmark for future optimization.

### 3.7 Impact of Saline Application on Signal Fidelity

Initial comparative analysis between the dry interface (Collection 1) and the saline-applied interface (Collection 2) at the dorsal wrist revealed no observable differences in ensemble-averaged waveforms (Supplementary Fig. S8). Because saline preparation provided no measurable uplift in signal fidelity to counter anticipated contact impedance, subsequent analyses were conducted using the dry interface data to better represent real-world consumer-wear conditions.

## 4 Discussion

This study presents the largest comparative evaluation of dual-modality hemodynamic sensing to date, analyzing over 150,000 cardiac cycles from 261 hypertensive and non-hypertensive individuals. Our primary finding is that IPG resolves a complex mixture of unimodal and arterial-dominant (A-line-like) morphologies. In contrast, PPG is limited by an information ceiling, consistently yielding broad, unimodal morphologies regardless of anatomical site or participant health status. While our cross-validated classification models achieved modest performance (a 14.3% improvement in AUC compared to a demographic-only model), the discovery and quantification of beat heterogeneity represents a critical milestone in cuffless BP research. To our knowledge, this is the first study to leverage morphological variance as a vital hemodynamic signature that is uniquely accessible through deep-tissue bioimpedance.

While previous IPG research established site-specific morphological variation in single-digit pilot cohorts [19] or relied on averaging individual variations into singular waveforms [22], these studies largely bypass the dorsal wrist - the primary anatomical site for consumer smartwatch integration. Furthermore, these small-scale trials (*N* = 4 to 15) often rely on acute interventions such as exercises - protocols where the confounding contribution of heart rate shifts to BP variance remains difficult to decouple[20, 21]. In contrast, our unsupervised analysis of 261 participants demonstrates that the inherent beat heterogeneity unique to IPG encodes vital pathological signatures that PPG’s superficial unimodal waveforms fail to resolve. By evaluating this diverse cohort across multiple sites - including the commercially-universal dorsal wrist - we reveal how physiological habitus significantly modulates the bioimpedance signal, showing a direct main effect of BP status on IPG amplitude (*p* = 0.024) and a BMI-dependent damping of the prevalence of steeper mode archetypes (*p* = 0.035).

The primary physiological rationale for the divergence between these two modalities lies in their respective sensing depths. It is well-established that PPG is restricted to the superficial dermal microvasculature, where arterial reflections are essentially low-pass filtered by the capillary beds [2, 3]. While competing models debate whether the PPG signal originates primarily from blood flow velocity or volumetric shifts in the capillary beds, researchers increasingly acknowledge that this shallow penetration depth may limit model performance [9]. Effectively, PPG captures a downstream hemodynamic consequence - localized tissue engorgement - rather than the primary arterial pressure wave itself. IPG, by contrast, leverages an electrical path-of-least-resistance that penetrates beyond the hypodermis to interrogate the deeper conductive volume of the limb. Our earlier work established that during temperature-mediated peripheral vasoconstriction, the PPG signal significantly diminishes or decouples from central changes, whereas the IPG signal remains robust [5]. This confirms that IPG bypasses superficial dermal artifacts to resolve signatures within larger-diameter blood vessels, or at least those unimpeded by peripheral vasoconstriction. Across every analytical framework employed in this study - from standard cross-correlation templates to GMM-separated profiles - the ability to resolve A-line-like waveforms remained a unique capability of IPG.

However, this observability was highly sensitive to anatomical localization; the ventral forearm exhibited a clear advantage over the dorsal wrist. Similarly, the ventral wrist measurements had a higher prevalence of the A-line-like mode compared to the dorsal site (Fig. 7). This is likely due to the physical proximity to the radial and ulnar arteries and the comparative lack of electrical shielding from the radius and ulna bones [27, 29]. These findings suggest a critical form-factor trade-off: while the dorsal-worn smartwatch remains the consumer standard, a smart band configured for ventral wrist contact may be required for more accurate BP monitoring. Alternatively, devices designed for the dorsal wrist may simply rotate to do spot-check measurements on the ventral side. This anatomical advantage provides a physiological rationale for the design of earlier commercial bioimpedance pioneers, such as the Jawbone UP3, which utilized a ventral electrode array to achieve superior accuracy in resting-state physiological tracking compared to its dorsal optical peers [23]. While maintaining consistent contact with rigid sensors on the ventral wrist presents mechanical challenges for active-wear (i.e., the display for commercial watches is on the dorsal side), our results indicate that this site offers the most high-fidelity window into deep-tissue signatures.

The impact of BMI on beat morphology is the secondary highlight of this work. We observed a clear morphological gradient where increasing BMI resulted in a dulling of the systolic upstroke and a corresponding elevation of the mid-diastolic pulse profile in IPG. Adipose tissue acts as both a mechanical barrier and an electrical insulator; because BMI correlates with wrist circumference (*r* = 0.53) [25, 26] and has statistical significance with artery depth [27], individuals with higher BMI face a sensing penalty where the desired arterial signal is buried deeper and insulated by thicker subcutaneous fat. Rather than a modality-specific limitation, this represents a fundamental biophysical challenge common to peripheral sensors, including PPG [24]. Since hypertension is a global health concern regardless of body habitus [28], these results underscore the need for BMI-aware models that can normalize for the insulating properties of adipose tissue.

Our classification result (AUC = 0.72), while promising, represents an incremental improvement upon demographic-only baselines and should be contextualized against earlier IPG pilot studies (*N ≤* 15) that report higher metrics by utilizing acute interventions and subject-specific calibration [19, 22, 20]. By opting for an uncalibrated, cross-sectional design in a diverse cohort of 261 participants, this work establishes a more realistic benchmark. It is yet to be determined whether these variations reflect transient autonomic shifts, localized vascular resonance, or additional confounders known to impact PPG such as pressure and posture [2, 9]. Future work should consider simultaneous IPG with A-line measurements or biophysics simulation to better understand the origin of beat heterogeneity for IPG. Furthermore, while our overall cohort was large, the Underweight BMI category suffered from a small sample size (*n ≤* 6), limiting our ability to generalize findings for that specific demographic. Another limitation of this study was that our hypertension classification model treated IPG and PPG independent of one another, whereas complementary incorporation of both signals might render different performance. Overall, the volumetric richness of IPG demands a shift from manual PWA to deep learning models, which we hypothesize will bridge the performance gap between wearable and clinical gold standards. Specifically, foundational models pre-trained on high-volume PPG databases might be leveraged to capture universal cardiac-driven temporal features, which could then be fine-tuned with IPG-specific deep tissue representations. Finally, our findings are limited by the stationary nature of the data collection protocol, which was conducted under controlled, resting-state conditions for a limited time duration (*≤* 10 minutes for each anatomical collection site per participant). Future work should evaluate the robustness of IPG in ambulatory settings across more dynamic, free-living environments.

This work demonstrates that bioimpedance offers a superior window into systemic hemodynamics that traditional optical sensors simply cannot reach. We have provided the largest dataset and a novel analytical pipeline to frame this problem, but much work remains to be done to fully crystallize the value proposition of IPG in the wearable market. We invite the research community to build upon these findings towards truly continuous, deep-tissue cardiovascular monitoring.

## Authorship Contribution Statement

S.T. and S.J. contributed equally to this work. S.T. and S.J. conceived and conceptualized the study, developed the methodology, performed data quality assurance, conducted data analysis, generated plots, and wrote the manuscript. A.P. provided supervision and suggested ideas for data analysis. A.D. provided ideas for data analysis. L.C. contributed to the conceptualization, methodology, conducted the study, and reviewed the manuscript. E.B. contributed to the conceptualization and conducted the study. J.W. provided ideas for data collection. D.M. provided supervision and ideas for data analysis. L.S. provided supervision and participated in discussions. S.S. provided funding and reviewed progress and results. S.P. provided funding. All authors reviewed and approved the final manuscript.

## Acknowledgements

This work was entirely supported by Google LLC. We would like to thank Paolo Di Achille, Ming-Zher Poh, Matthew Thompson, and Tracy Geist for their time and feedback during the project. We would also like to thank Eric Mockensturm for his suggestion to lean into the BMI analysis, and Conor Heneghan for reviewing this manuscript.

## Data Availability

Data and code used in this study will be made publicly available upon the acceptance of this manuscript.

## 5 Supplementary Figures

**Supplementary Figure S1:**
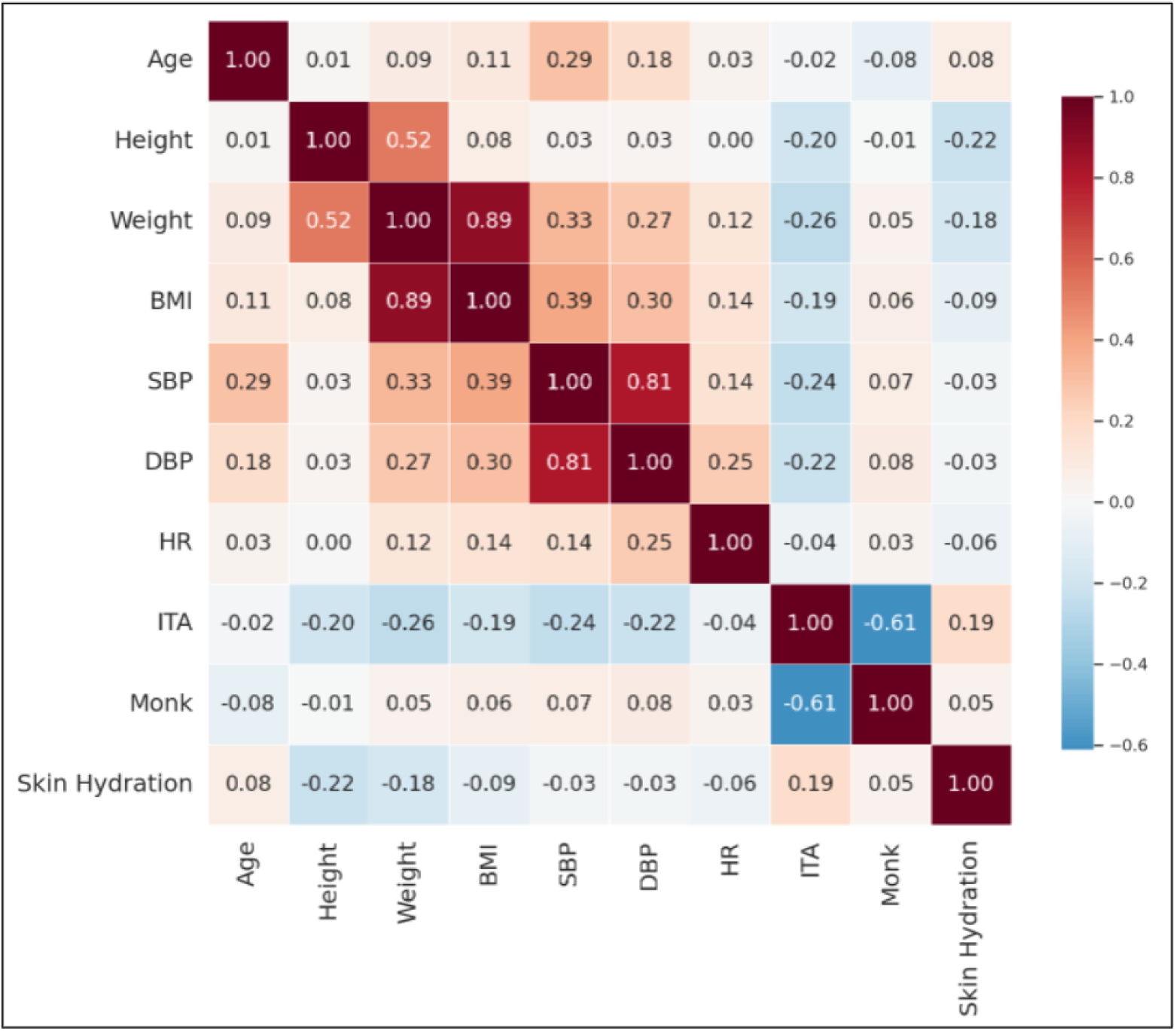
Correlation matrix of baseline demographic and clinical variables. Heatmap illustrates Pearson correlation coefficients between key physiological, demographic, and sensor-relevant characteristics across the entire study cohort (N = 261). Strong expected collinearities are observed between weight and BMI (r = 0.89) as well as systolic and diastolic blood pressure (r = 0.81). Subjective Monk Skin Tone and objective melanin content (Colorimeter ITA°) demonstrate a moderate negative correlation (r = -0.61).

**Supplementary Figure S2:**
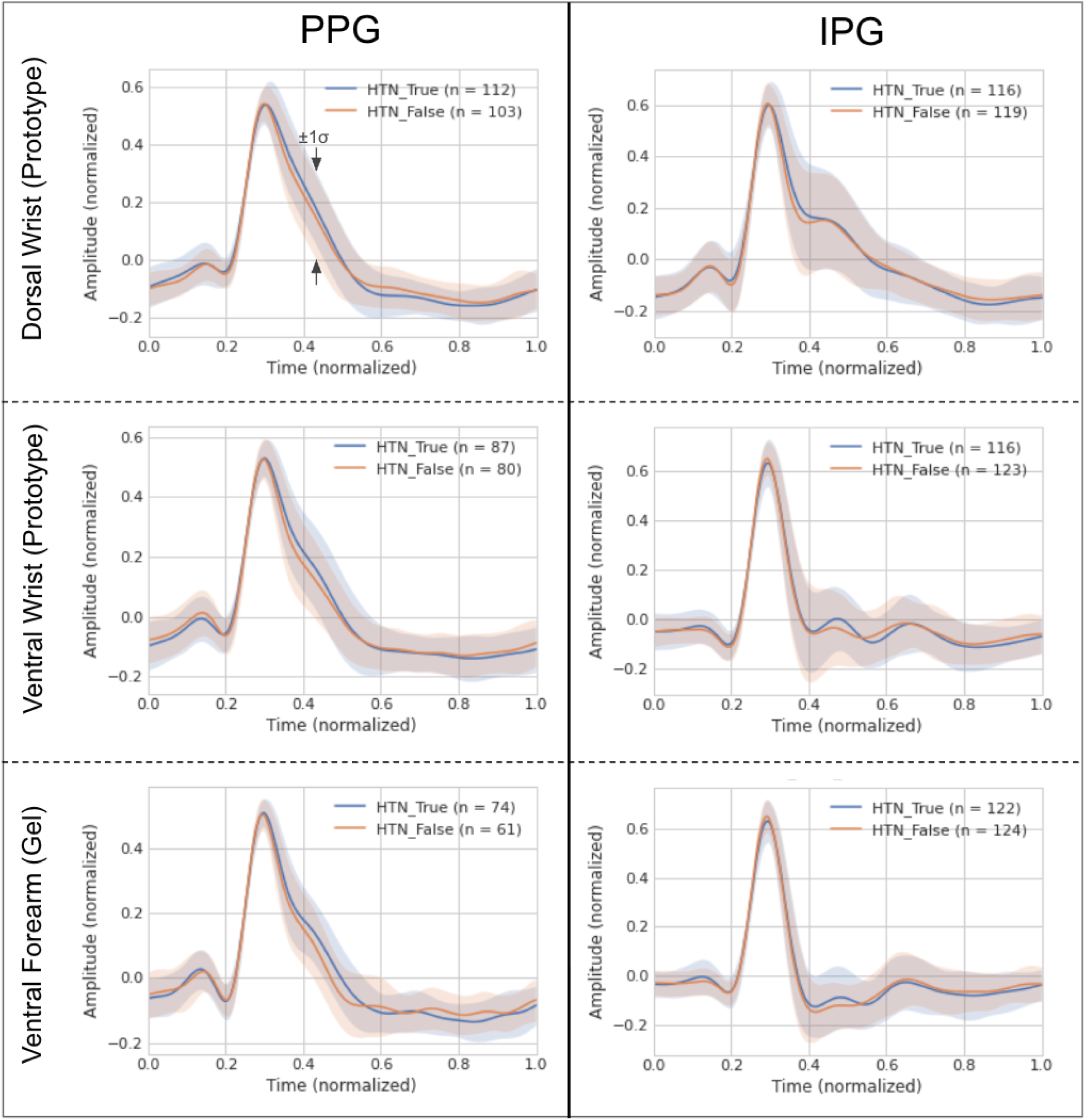
First time derivatives of comparative PPG and IPG waveforms. Ensemble-averaged first time derivatives (mean ± 1 standard deviation) for PPG (left column) and IPG (right column) signals, stratified by non-hypertensive (HTN_False, orange) and hypertensive (HTN_True, blue) cohorts. Data are evaluated across the dorsal wrist (top), ventral wrist (middle), and ventral forearm reference site (bottom). The IPG velocity profiles exhibit sharper, more complex multi-phasic inflection points - particularly during the diastolic decay - contrasting with the heavily smoothed, low-pass-filtered appearance of the PPG derivatives.

**Supplementary Figure S3:**
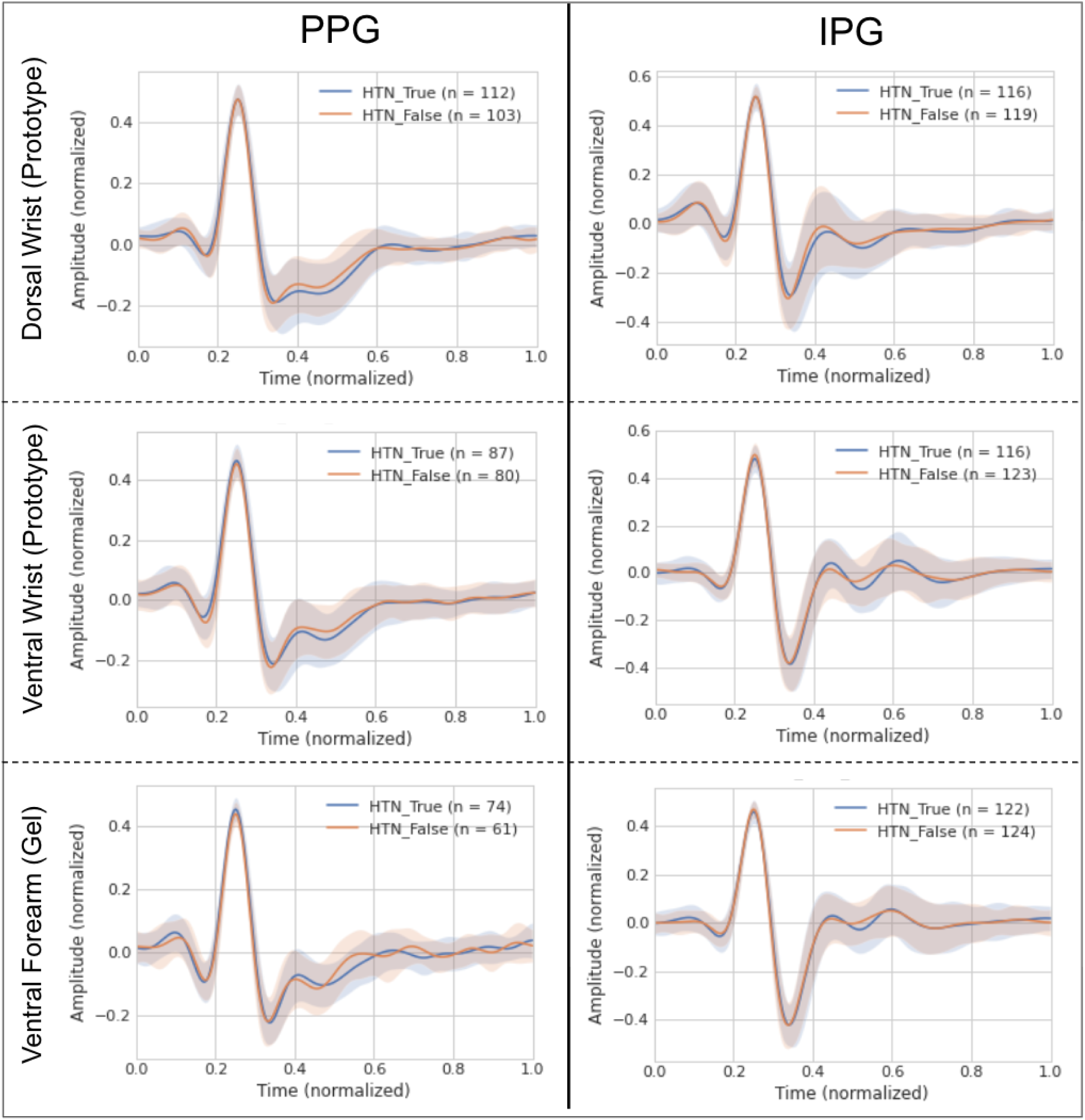
Second time derivatives of comparative PPG and IPG waveforms. Ensemble-averaged second time derivatives (mean ± 1 standard deviation) for PPG (left column) and IPG (right column) signals, stratified by non-hypertensive (HTN_False, orange) and hypertensive (HTN_True, blue) cohorts across three anatomical sites. The acceleration profiles of the IPG signals highlight highly localized hemodynamic events, rendering subtle fiducial landmarks (such as the dicrotic notch and secondary diastolic reflections) far more resolvable than in the spatially coincident PPG signals.

**Supplementary Figure S4:**
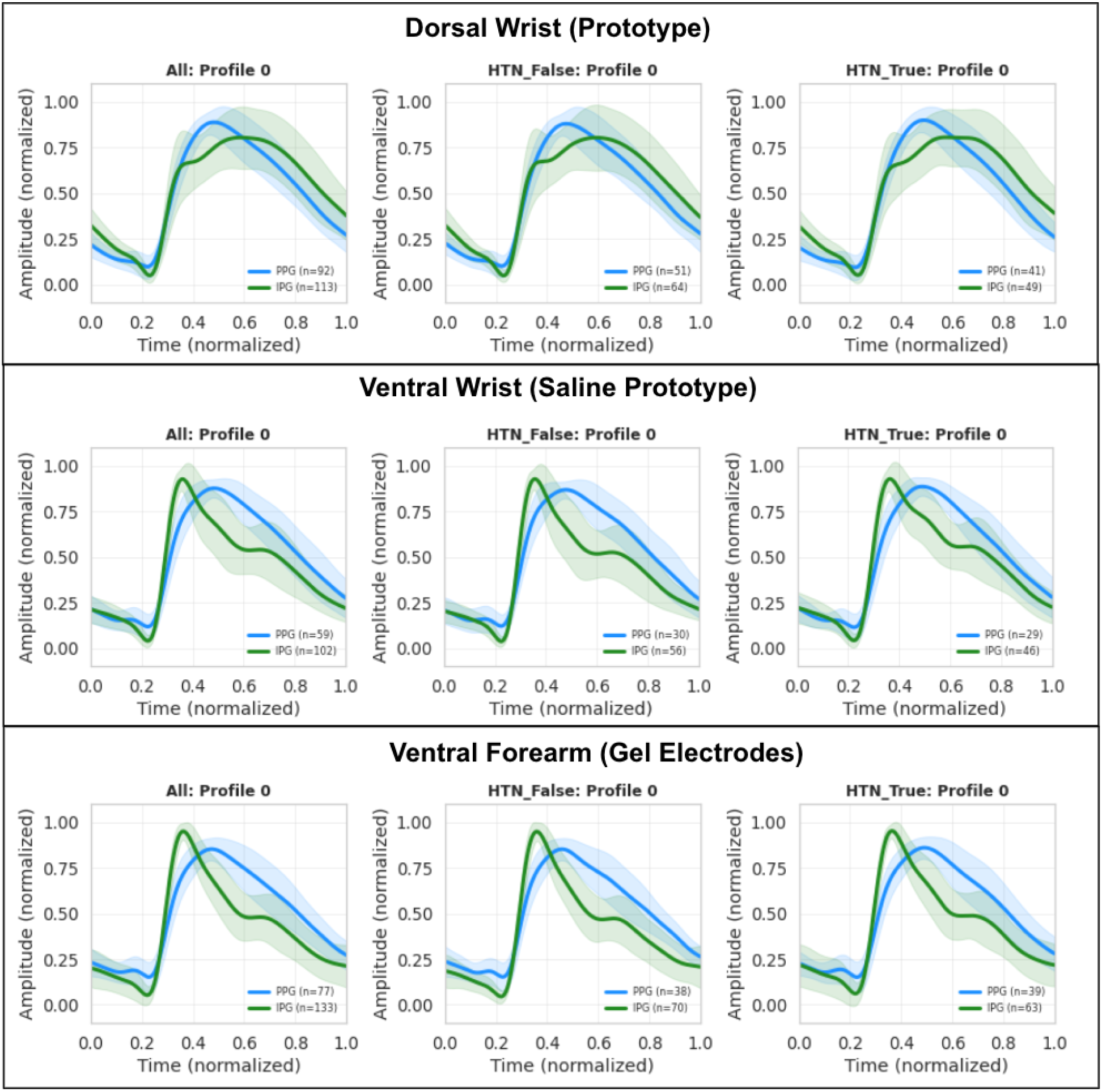
Morphological divergence between hypertensive and non-hypertensive cohorts within IPG Profile 0. Ensemble-averaged waveforms (mean *±*1 standard deviation) isolating participants classified into the A-line-like Profile 0 via Gaussian Mixture Modeling (restricted to GMM confidence level *>* 0.85). While PPG (blue) remains morphologically invariant between cohorts, IPG (green) captures distinct pathological signatures in the hypertensive group. Specifically, at the dorsal and ventral wrist sites, hypertensive participants exhibit a pronounced mid-diastolic plateau and elevated tidal wave components compared to the more rapid diastolic decay observed in non-hypertensive participants.

**Supplementary Figure S5:**
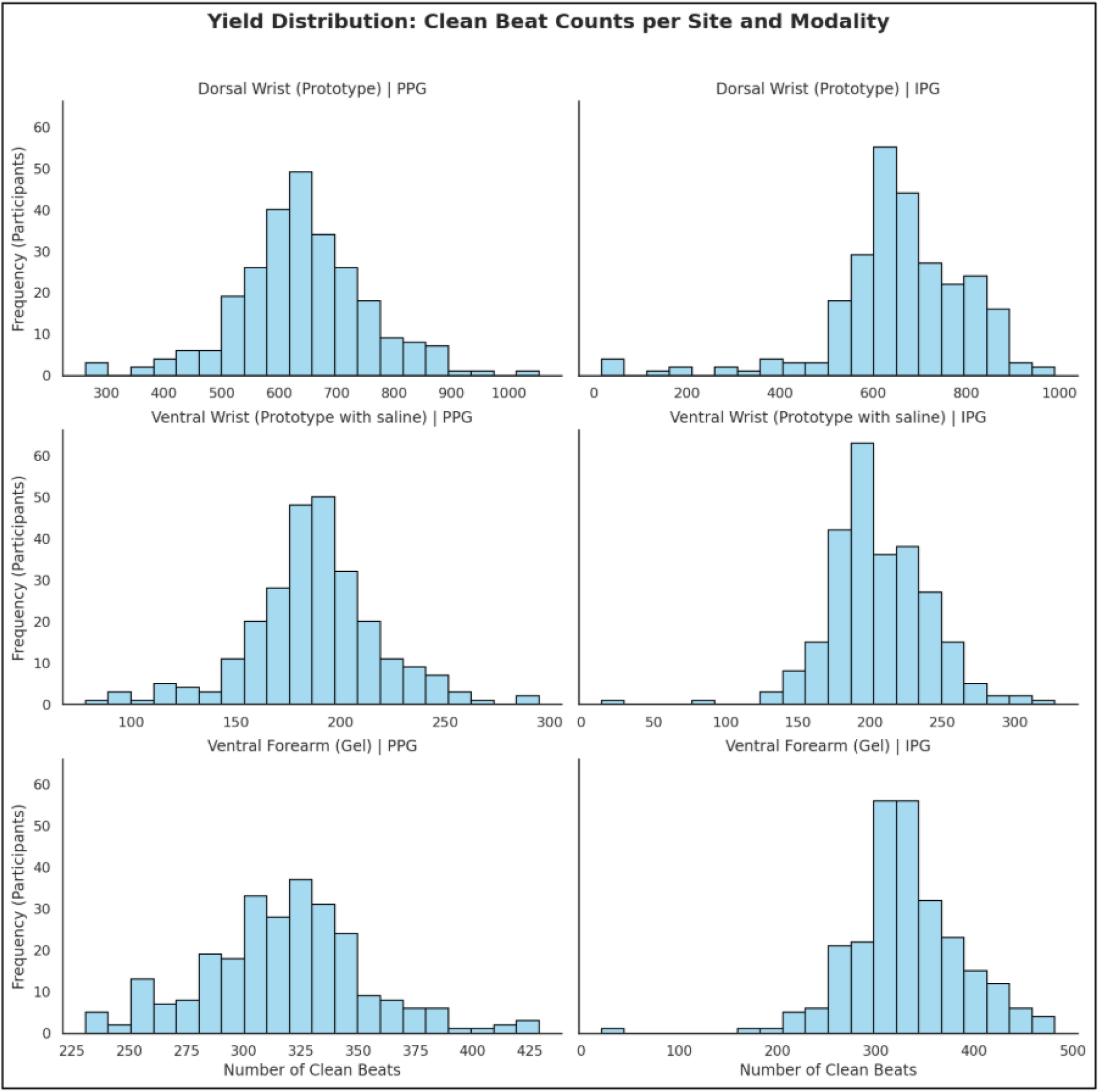
Distribution of high-quality cardiac cycle yield across sensing modalities and anatomical sites. Histograms illustrate the frequency of clean beats retained per participant. Data are stratified by modality (PPG left column, IPG right column) across three measurement locations: dorsal wrist prototype (top), ventral wrist prototype with saline (middle), and the ventral forearm (gel) (bottom).

**Supplementary Figure S6:**
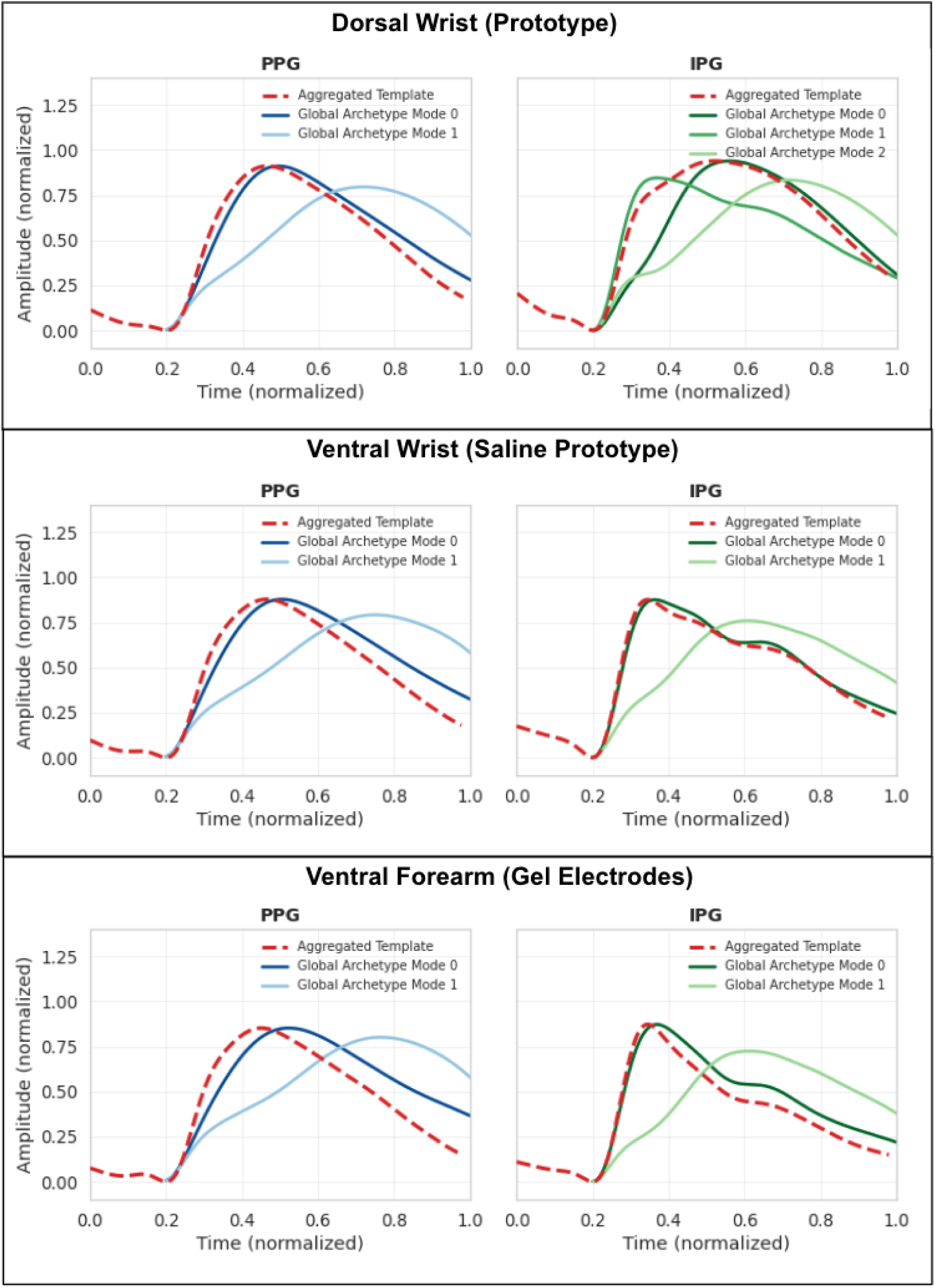
Comparison of ensemble-averaged templates against unsupervised global archetypes. Representative waveforms are shown across three anatomical sites for PPG (left) and IPG (right). The population-wide ensemble average, derived via standard cross-correlation (aggregated template), is represented by the red dashed line. The distinct morphological archetypes identified through unsupervised PCA and K-means clustering are represented by the solid lines. This visualization demonstrates how traditional single-template averaging obscures the complex hemodynamic signatures captured by IPG.

**Supplementary Figure S7:**
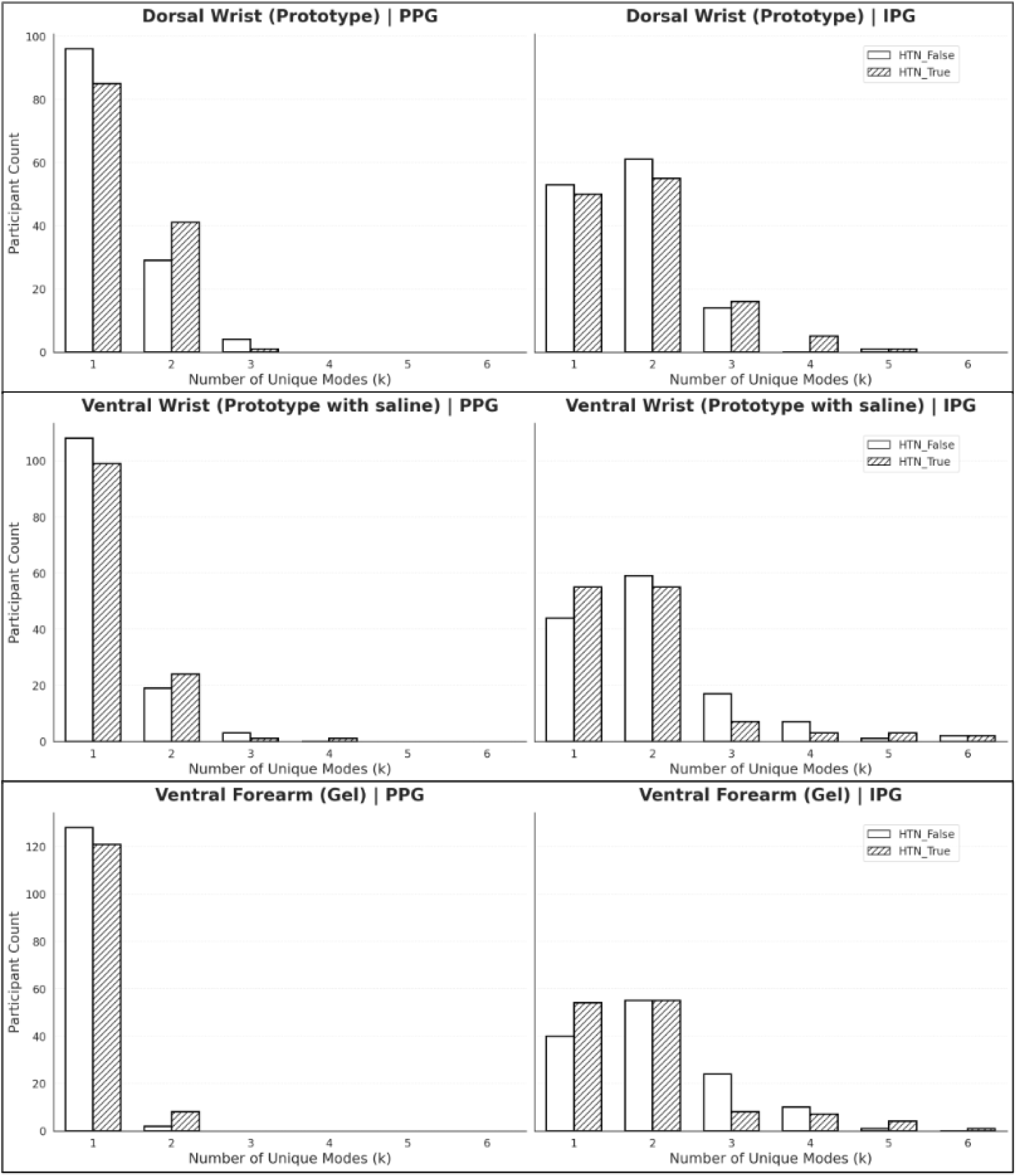
Distribution of intra-individual morphological complexity across modalities. Histograms detailing the number of unique morphological modes (k) identified per participant using an individualized K-means clustering pipeline (Silhouette threshold = 0.40). Data are stratified by non-hypertensive (HTN_False, white) and hypertensive (HTN_True, hatched) cohorts for PPG (left column) and IPG (right column). Across all anatomical sites, PPG is heavily skewed toward a singular morphological mode (k = 1). Conversely, a substantial proportion of the cohort exhibits multiple distinct beat archetypes (k *≥* 2) when evaluated with IPG.

**Supplementary Figure S8:**
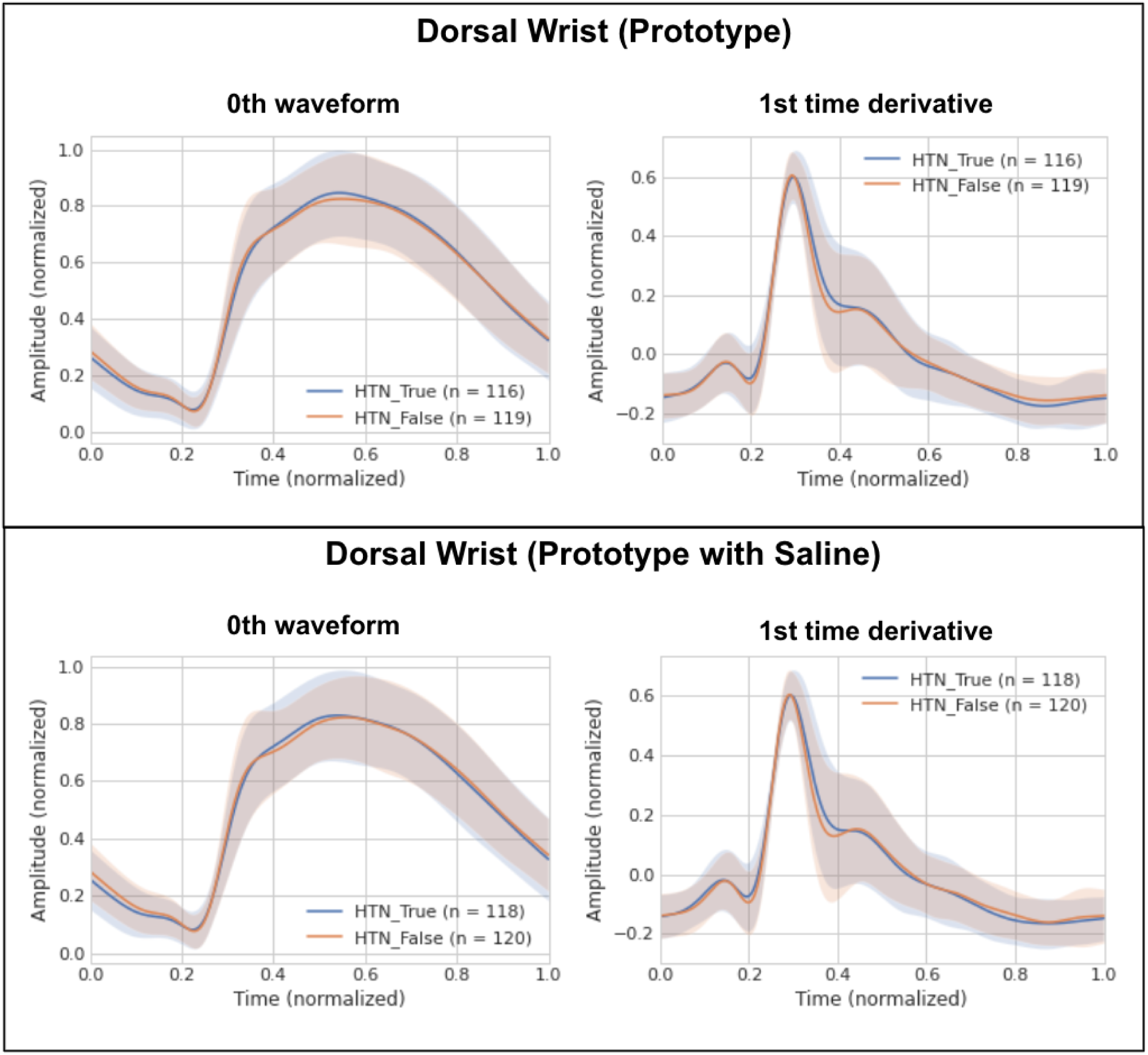
Comparative analysis of IPG signal morphology across dry and saline skin-sensor interfaces. Ensemble-averaged waveforms (left column) and their corresponding first time derivatives (right column) for IPG signals acquired at the dorsal wrist prototype. Data are compared between standard dry-electrode conditions (top row) and saline-prepared conditions (bottom row) to assess the impact of contact impedance. Signals are stratified by hypertension status. No observable morphological degradation or signal fidelity loss was noted under the dry-interface condition.

## 6 Supplementary Table

**Supplementary Table 1:** Welch’s ANOVA for *A*_sp_ across BMI Categories (Training Set)

| Sensor | Measurement | F-value | p-value |
| --- | --- | --- | --- |
| PPG | Dorsal Wrist (Prototype) | 4.817 | 0.010* |
|  | Ventral Wrist (Prototype) | 1.110 | 0.334 |
|  | Ventral Forearm (Gel) | 1.926 | 0.153 |
| IPG | Dorsal Wrist (Prototype) | 5.059 | 0.008* |
|  | Ventral Wrist (Prototype) | 0.078 | 0.925 |
|  | Ventral Forearm (Gel) | 7.639 | < 0.001* |

**Supplementary Table 2:** Games-Howell Post-hoc Test for Asp across BMI Categories (Training Set)

| Sensor | Measurement | (I) Region | (J) Region | Mean difference (I-J) | p-value |
| --- | --- | --- | --- | --- | --- |
| PPG | Dorsal Wrist (Prototype) | Overweight | Underweight/Healthy | -0.0118 | 0.121 |
|  |  | Overweight | Obese | 0.0062 | 0.232 |
|  |  | Underweight/Healthy | Obese | 0.0180 | 0.009* |
|  | Ventral Wrist (Prototype) | Overweight | Underweight/Healthy | -0.0040 | 0.583 |
|  |  | Overweight | Obese | 0.0021 | 0.837 |
|  |  | Underweight/Healthy | Obese | 0.0062 | 0.303 |
|  | Ventral Forearm (Gel) | Overweight | Underweight/Healthy | -0.0004 | 0.991 |
|  |  | Overweight | Obese | -0.0053 | 0.141 |
|  |  | Underweight/Healthy | Obese | -0.0050 | 0.311 |
| IPG | Dorsal Wrist (Prototype) | Overweight | Underweight/Healthy | 0.0159 | 0.021* |
|  |  | Overweight | Obese | 0.0058 | 0.698 |
|  |  | Underweight/Healthy | Obese | -0.0101 | 0.115 |
|  | Ventral Wrist (Prototype) | Overweight | Underweight/Healthy | -0.0014 | 0.930 |
|  |  | Overweight | Obese | -0.0002 | 0.998 |
|  |  | Underweight/Healthy | Obese | 0.0012 | 0.946 |
|  | Ventral Forearm (Gel) | Overweight | Underweight/Healthy | -0.0010 | 0.657 |
|  |  | Overweight | Obese | 0.0027 | 0.038* |
|  |  | Underweight/Healthy | Obese | 0.0037 | < 0.001* |

**Supplementary Table 3:**
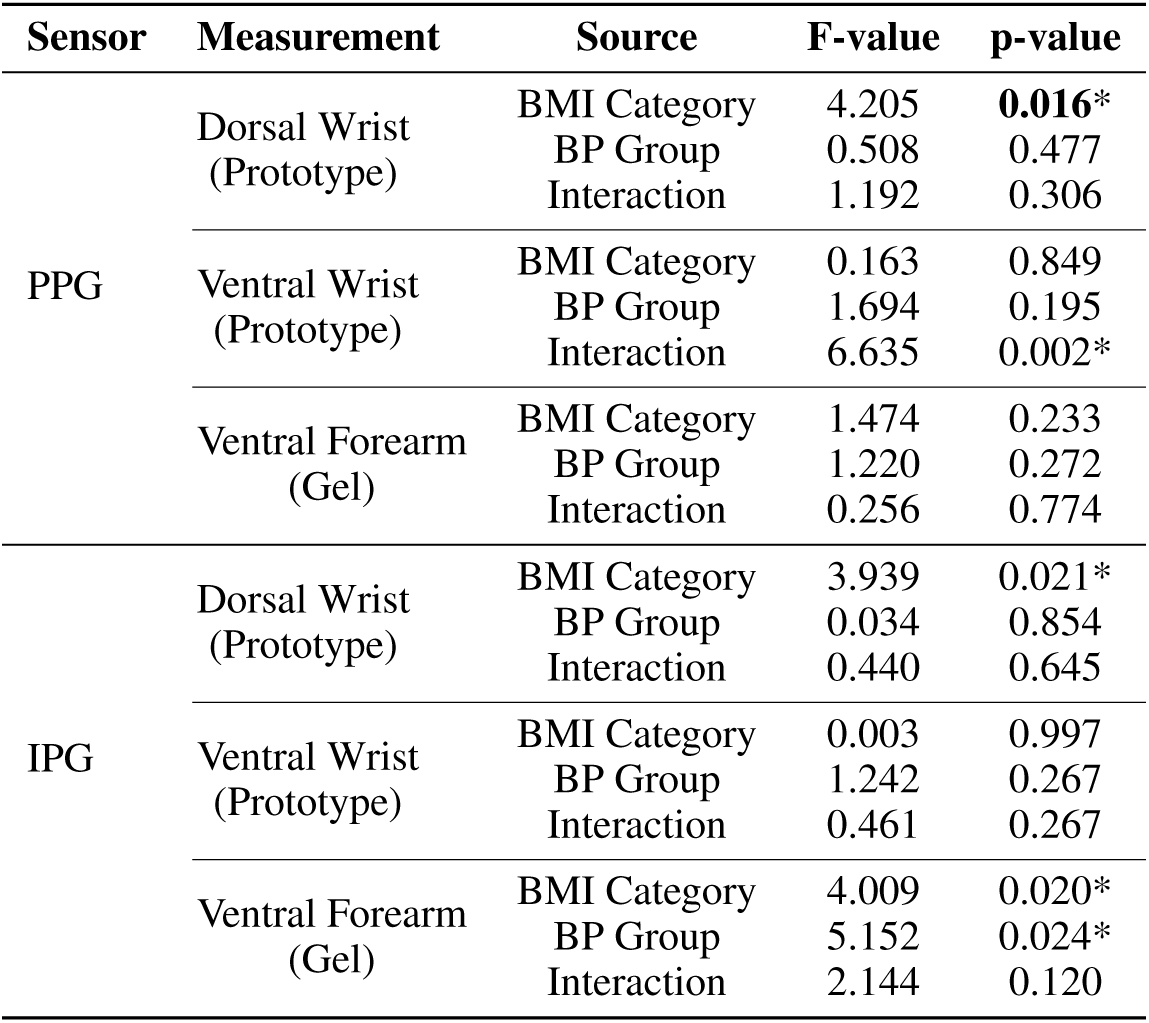
Two-Way ANOVA for Asp across BMI Categories and BP Groups (Training Set)

| Sensor | Measurement | Source | F-value | p-value |
| --- | --- | --- | --- | --- |
| PPG | Dorsal Wrist (Prototype) | BMI Category | 4.205 | **0.016\*** |
|  |  | BP Group | 0.508 | 0.477 |
|  |  | Interaction | 1.192 | 0.306 |
|  | Ventral Wrist (Prototype) | BMI Category | 0.163 | 0.849 |
|  |  | BP Group | 1.694 | 0.195 |
|  |  | Interaction | 6.635 | 0.002* |
|  | Ventral Forearm (Gel) | BMI Category | 1.474 | 0.233 |
|  |  | BP Group | 1.220 | 0.272 |
|  |  | Interaction | 0.256 | 0.774 |
| IPG | Dorsal Wrist (Prototype) | BMI Category | 3.939 | 0.021* |
|  |  | BP Group | 0.034 | 0.854 |
|  |  | Interaction | 0.440 | 0.645 |
|  | Ventral Wrist (Prototype) | BMI Category | 0.003 | 0.997 |
|  |  | BP Group | 1.242 | 0.267 |
|  |  | Interaction | 0.461 | 0.267 |
|  | Ventral Forearm (Gel) | BMI Category | 4.009 | 0.020* |
|  |  | BP Group | 5.152 | 0.024* |
|  |  | Interaction | 2.144 | 0.120 |

**Supplementary Table 4:** Games-Howell Post-hoc Test for Asp across BMI Categories for each BP group (Training Set)

| Sensor | Measurement | BP group | (I) Region | (J) Region | Mean difference (I-J) | p-value |
| --- | --- | --- | --- | --- | --- | --- |
| PPG | Ventral Wrist (Prototype) | HTN_False | Overweight | Obese | 0.0139 | 0.020* |
|  |  | HTN_True | Obese | Overweight | 0.0149 | 0.009* |

**Supplementary Table 5:** Results of the Generalized Linear Model for the prevalence of the steepest pulse morphology. Wald chi-squared test applied to (Training Set)

| Sensor | Measurement | Steepest Mode | HTN_True vs HTN_False p-value | Underweight/Healthy vs Overweight p-value | Normal vs Obese p-value |
| --- | --- | --- | --- | --- | --- |
| IPG | Dorsal Wrist (Prototype) | Mode 1 | 0.661 | 0.245 | 0.072 |
|  | Ventral Wrist (Prototype) | Mode 2 | 0.174 | 0.687 | 0.741 |
|  | Ventral Forearm (Gel) | Mode 1 | 0.154 | 0.512 | 0.035* |
| PPG | Dorsal Wrist (Prototype) | Mode 1 | 0.694 | 0.261 | 0.121 |
|  | Ventral Wrist (Prototype) | Mode 2 | 0.802 | 0.819 | 0.658 |
|  | Ventral Forearm (Gel) | Mode 1 | 0.911 | 0.681 | 0.303 |

